# Genomic ascertainment of *PALB2*-related cancer predisposition

**DOI:** 10.64898/2026.04.03.26349984

**Authors:** Douglas R. Stewart, Jung Kim, Jeremy S. Haley, Jiang Li, Michael R. Sargen, Hyokyoung Hong, Marc Tischkowitz, Lisa J. McReynolds, David J. Carey

## Abstract

**PURPOSE:** To evaluate cancer risk, age-specific penetrance, and mortality associated with heterozygous pathogenic or likely pathogenic (P/LP) germline *PALB2* variants identified through genomic ascertainment and to assess modification by family history of cancer.

**PATIENTS AND METHODS:** We conducted a case-control study in two large population-based adult cohorts: the UK Biobank (n=469,580) and Geisinger MyCode (n=167,050). Individuals with heterozygous *PALB2* P/LP variants were identified via exome sequencing and compared with non-carriers. Cancer diagnoses and vital status were obtained from linked registry and electronic health record data. We used multivariable logistic regression to estimate odds ratios (ORs) for cancer outcomes and Cox proportional hazards models to estimate hazard ratios (HRs) for all-cause mortality. Age-specific cumulative incidence (penetrance) was estimated using Kaplan–Meier methods. Models were adjusted for birth year, sex (when applicable), smoking status, and body mass index; stratified analyses assessed modification by family history of cancer.

**RESULTS:** *PALB2* P/LP variant prevalence was 1:571 in UK Biobank and 1:940 in MyCode, with the higher prevalence in the UK cohort driven by the *PALB2* p.Trp1038Ter founder variant. Compared with non-carriers, heterozygotes had significantly increased odds of any cancer, female breast cancer, pancreatic cancer, and cancers of ill-defined or secondary sites in both cohorts (P < 0.01). Adjusted hazard ratios for any cancer and female breast cancer ranged from 1.7 to 3.6. All-cause mortality was increased among *PALB2*-heterozygotes (HR 1.61–1.67), and survival after cancer diagnosis was reduced. Family history further modified cancer risk.

**CONCLUSION:** Genomic ascertainment of *PALB2*-heterozygotes identifies elevated risk for multiple cancers and increased mortality, although risks were lower than estimates from familial ascertainment. These findings inform risk management for individuals identified through genomic screening.

## Introduction

The Partner and Localizer of BRCA2 (PALB2) interacts with both BRCA2 and BRCA1 to ensure genomic stability via homologous recombination repair of DNA double-stranded breaks.^1^ Biallelic germline loss-of-function variants in *PALB2* (also known as *FANCN*) lead to Fanconi anemia.^2^ Elevated cancer risk in female and male breast,^3,4^ pancreas^5^ and ovarian^6^ cancer in individuals harboring a pathogenic variant in *PALB2* is well-established, with no consistent evidence for increased risk of other cancers. *PALB2* is routinely included on multigene panel tests^7^ and is on the American College of Medical Genetics (ACMG) Secondary Findings list^8^.

Cancer risk arising from a germline pathogenic *PALB2* variant, especially ascertained through a personal or family history of cancer, has been well-studied. Genomic ascertainment is the reverse of traditional phenotypic ascertainment insofar as cases are defined by identifying germline variation of interest and then determining phenotype status^9^, an approach that can result in lower risk estimates.^10^ What is not known is the magnitude of cancer risk when *PALB2* pathogenic variation is genomically ascertained, for example through opportunistic screening (e.g., secondary findings) or incidentally, nor how this risk is influenced by a family history of cancer. In addition, overall mortality and cancer-associated mortality from genomic ascertainment are unknown.

In this case-control analysis, we used genomic ascertainment in two population-based cohorts (UK Biobank (UKBB) and Geisinger MyCode) to estimate the prevalence of *PALB2*-heterozygotes and to evaluate age-specific penetrance, cancer risks, and survival outcomes among carriers compared with controls (non-carriers).

## Materials and Methods

### Cohorts and relatedness

UKBB germline variants were obtained from field 23157, population level exome OQFE variants, and pVCF format, final exome release with 351 individual withdrawal list (downloaded 07/02/2024). UKBB exome sequencing has been previously described.^11^ Human subjects’ protection and review was through the North West Multi-centre Research Ethics Committee. The number of unrelated participants was determined by the R package ukbtools, “ukb_gen_related_with_data” function.

The Geisinger MyCode cohort has been previously described^12^ and approved by the Geisinger Institutional Review Board; DNA samples were exome-sequenced by the Regeneron Genetics Center as previously described.^13^ In this study, we included only individuals who are ≥18 years of age (n=167,050). To remove related individuals while maintaining the largest possible cohort, kinship pairs up to third-degree relatives (minimum PI_HAT = 0.1875) were used to create a graph of all relatives.

### Variant filtering and PALB2 pathogenicity classification

All germline variants were filtered on the following quality metrics excluding those with genotype quality < 30, total read depth < 5, Allelic Balance of Heterozygotes (ABHet) < 0.3 or > 0.7. Variants that passed quality metrics were annotated using AutoGVP^14^ v1.0.1 with ClinVar database retrieved on 07/01/2024 and InterVar v2.1.3.^15^ *PALB2* pathogenic (P) or likely pathogenic (LP) from AutoGVP classifications were reviewed by a medical geneticist (MT). Individuals with a *PALB2* P/LP variant are hereafter “*PALB2-*heterozygote.” Controls were individuals who did not harbor a *PALB2* P/LP variant.

### Cancer phenotype and vital status query

Demographic data (age, sex, body mass index (BMI), alcohol consumption, smoking history, and race) were obtained for both heterozygotes and controls and comparisons were completed using Student’s t-test and Fisher’s exact test. In the UKBB, cancer and vital status phenotypes were obtained from cancer registry (field ID 40006, 40013), hospital in-patient EHR (field ID 41270, 41271), and death registry (field ID 40001). In MyCode, cancer and vital status phenotypes were obtained from the Geisinger Cancer Registry and EHR.

### Power to detect predisposition to common and rare cancers in MyCode and UKBB

Power to detect associations between *PALB2-*heterozygotes and cancer outcomes was evaluated using the method of Chow et al. under a cohort study framework.^16^ **Supplemental Figures 1 and 2** show power curves for a range of assumed true odds ratios and cancer event rates in MyCode and UKBB, respectively, based on the cohort-specific *PALB2*-heterozygote frequencies in **Supplemental Table 1**. Both cohorts had ∼90% power to detect associations for cancers with event rates ≥5% and true odds ratios >2. For cancers with event rates ≥1%, UKBB had ≥80% power to detect an odds ratio >2, while MyCode had ≥80% power to detect an odds ratio >3. **Supplemental Figure 3** shows power estimates in UKBB using the frequency of the *PALB2* p.Trp1038Ter founder variant. **Supplemental Figure 4** shows analogous estimates using the frequency of *PALB2*-heterozygotes excluding p.Trp1038Ter.

## Statistical analyses

### Cancer association analyses

Associations between *PALB2* heterozygosity and cancer outcomes were evaluated using multivariable logistic regression, estimating odds ratios (ORs) and 95% confidence intervals (CIs). These models assessed the association between *PALB2* carrier status and the presence of a recorded cancer diagnosis by the end of available follow-up, adjusted for year of birth, sex (when applicable), smoking status, and body mass index. Analyses of sex-specific cancers were restricted to the relevant sex. Cancer outcomes were defined using ICD-10 codes and grouped by organ system.

### Time-to-event and penetrance analyses

Time-to-event outcomes, including time to first cancer diagnosis and all-cause mortality, were analyzed using Cox proportional hazards models, estimating hazard ratios (HRs) and 95% CIs. Age was used as the underlying time scale, with individuals entering the risk set at age of cohort enrollment and followed until the event of interest, death, or censoring at last known follow-up.

Kaplan-Meier curves were used to visualize time-to-event distributions and to estimate age-specific cumulative incidence (penetrance) among *PALB2*-heterozygotes. These penetrance estimates represent cohort-specific cumulative incidence conditional on survival to enrollment age and observation through follow-up.

### Family history analyses

In UKBB, family history of cancer was assessed using data fields 20110 (“illness of mother”) and 20111 (“illnesses of siblings”). Effect modification by family history was evaluated through stratified analyses.

### Software

All statistical analyses were performed in R version 4.3.2.

## Results

### Prevalence and demographics of PALB2-heterozygotes in MyCode and UKBB

**Supplemental Table 1** shows the prevalence of *PALB2*-heterozygotes and p.Trp1038Ter heterozygotes in both cohorts. In UKBB, the greater prevalence of *PALB2*-heterozygotes was driven by an excess of p.Trp1038Ter, a common variant in the British Isles as per the Regeneron Million Exome Variant Browser^17^ (version 1.1.3: https://rgc-research.regeneron.com/me/rsid/rs180177132). **Supplemental Table 2** has details on the variants. **Supplemental Table 3** lists demographic and covariate data in *PALB2*-heterozygotes and controls.

### Significant excess risk for all cancers, breast, digestive organs and ill-defined, other and secondary sites in PALB2-heterozygotes in both MyCode and UKBB

**Figure 1** displays the odds ratio of *PALB2*-heterozygotes for all significant organ system groupings of cancer ICD codes. In both cohorts, there was a significantly elevated odds of any cancer, malignant neoplasms of digestive organs (C15-C26), breast cancer (C50) and malignant neoplasms of ill-defined, other secondary and unspecified sites (C76-C80). In UKBB, there was one *PALB2-*heterozygote (0.26%) and 151 controls (0.07%) with a male breast cancer diagnosis; in MyCode, there were no male *PALB2*-heterozygotes with breast cancer. In MyCode, there was a significantly elevated odds of malignant and secondary neuroendocrine tumors, however the number of observations was modest (<5/cohort). In UKBB there was one *PALB2*-heterozygote and 225 controls with a neuroendocrine histology code (8013/3, 8246/3, 8574/3) with a nominal P = 0.048. **Supplemental Figure 5** includes all organ systems; the signal in the digestive organs in both cohorts arises from neoplasms of the pancreas. In UKBB there was a significant excess of melanoma and other malignant neoplasms of skin (C43-C44) and malignant neoplasms of female genital organs (C51-C58) but this was not observed in MyCode. **Supplemental Figure 5** shows that the skin and female genital organ neoplasm signals were driven by both melanoma and non-melanoma skin cancers and ovarian tumors, respectively.

**Figure 1.**
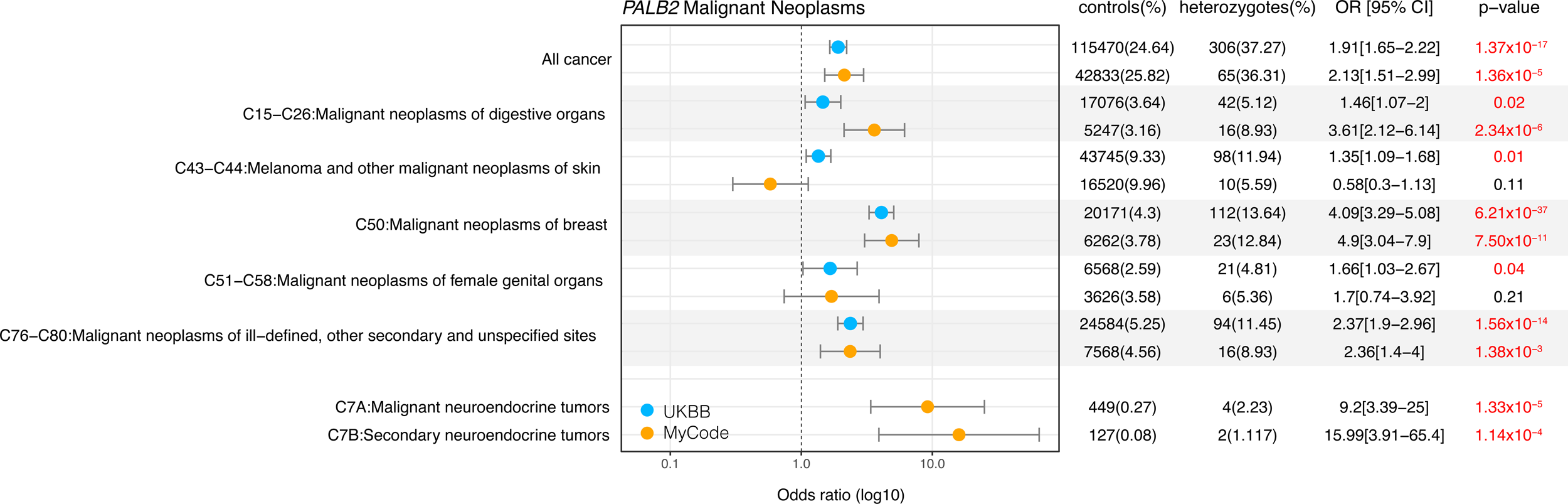
Odds ratio for *PALB2*-heterozygotes for organ system groupings of cancer ICD codes with a significant excess of risk in MyCode and UKBB. CI: 95% confidence interval; OR: odds ratio

**Supplemental Figure 6** displays the odds ratio of pathogenic *PALB2*-heterozygotes for carcinoma in situ codes. In both cohorts, there was a significantly elevated odds of carcinoma in situ of breast (D05); of these, D05.1 (ductal carcinoma in situ of breast (DCIS)), of which there were 20 in UKBB (OR: 3.29 [2.10-5.15] P = 2.21 x 10^-7^ (adjusted for sex, BMI, year of birth, smoking)), and 3 in Geisinger (OR: 2.86 [0.90-9.05] P = 0.074). In UKBB only, there was excess odds for carcinoma in situ of skin.

**Supplemental Figure 7** displays the odds ratio of *PALB2*-heterozygotes for benign tumor codes. In UKBB (only), there was a significant excess risk of benign neoplasms of skin (D17, D18, D22, D23) tumors and a decreased risk of benign neoplasm of female genital organs (D25-D27). In MyCode (only) there was elevated odds for benign neuroendocrine tumors (D3A) and benign neoplasm of mesothelial and soft tissue (D19-D21). **Supplemental Figure 8** shows the odds ratio of *PALB2*-heterozygotes for tumors of uncertain behavior in both cohorts.

### Time-to-all-cancer and time-to-breast cancer significantly younger in PALB2-heterozygotes vs controls in both cohorts and is influenced by family history of cancer

Compared to controls, time-to-all-cancer in *PALB2*-heterozygotes had significantly elevated hazard of any cancer in both MyCode (adjusted HR: 1.74 [95% CI 1.38-2.24], P = 5.94 x 10^-6^) and UKBB (adjusted HR 1.74 [95% CI 1.55-1.96], P-value: 4.54 x 10^-20^) (**Figure 2). Supplemental Table 4** shows the per-decade cumulative incidence of cancer stratified by sex for both cohorts. In both cohorts, excess cancer risk appears around age 40 years; by age 80 years, 65.8% (95% CI 51.5–75.9) and 44.8% (95% CI 40.1-49.2) of *PALB2*-heterozygotes had a cancer diagnosis in MyCode and UKBB, respectively. Compared to controls, *PALB2*-heterozygote women had a significantly elevated hazard of breast cancer in both MyCode (adjusted HR: 1.85 [95% CI 1.35-2.54], P = 1.54 x 10^-4^) and UKBB (adjusted HR 3.56 [95% CI 2.92-4.3], P = 9.27 x 10^-36^) (**Figure 3). Supplemental Table 5** shows the per-decade cumulative incidence of breast cancer for both cohorts. In both cohorts, excess breast cancer incidence appears around age 40 years; by age 80 years, 37.9% (95% CI 21.2–51.0) and 27.4% (95% CI 22.1–32.4) of female *PALB2*-heterozygotes had a cancer diagnosis in MyCode and UKBB, respectively. **Figure 4** shows time-to-breast-cancer in *PALB2*-heterozygotes with and without family history of breast cancer in UKBB; female *PALB2*-heterozygotes with a family history of breast cancer had a non-significantly increased risk of developing breast cancer compared with female *PALB2*-heterozygotes without a family history of breast cancer. (Similar family history data was not available from the MyCode cohort.) **Supplemental Table 6** shows the per-decade cumulative incidence of breast cancer for *PALB2*-heterozygotes with and without a family history of breast cancer.

**Figure 2.**
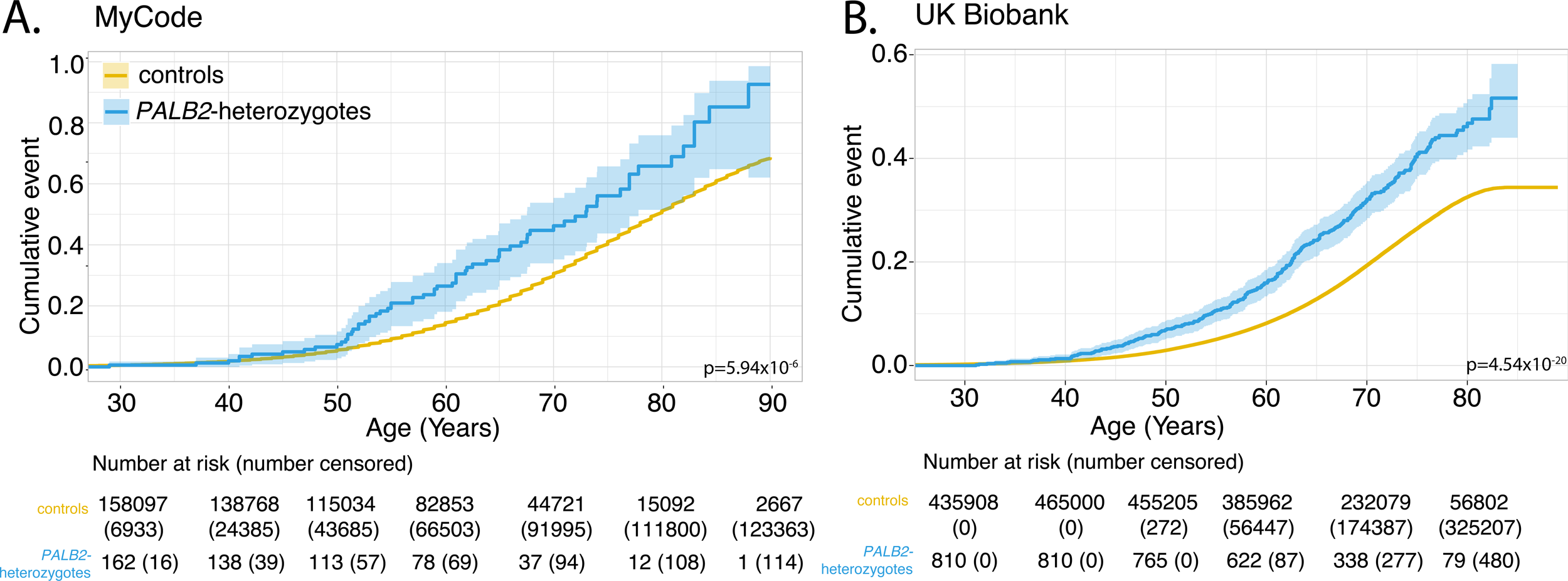
Time-to-all-cancer in MyCode (Panel A) and UKBB (Panel B). See also **Supplemental Table 4**.

**Figure 3.**
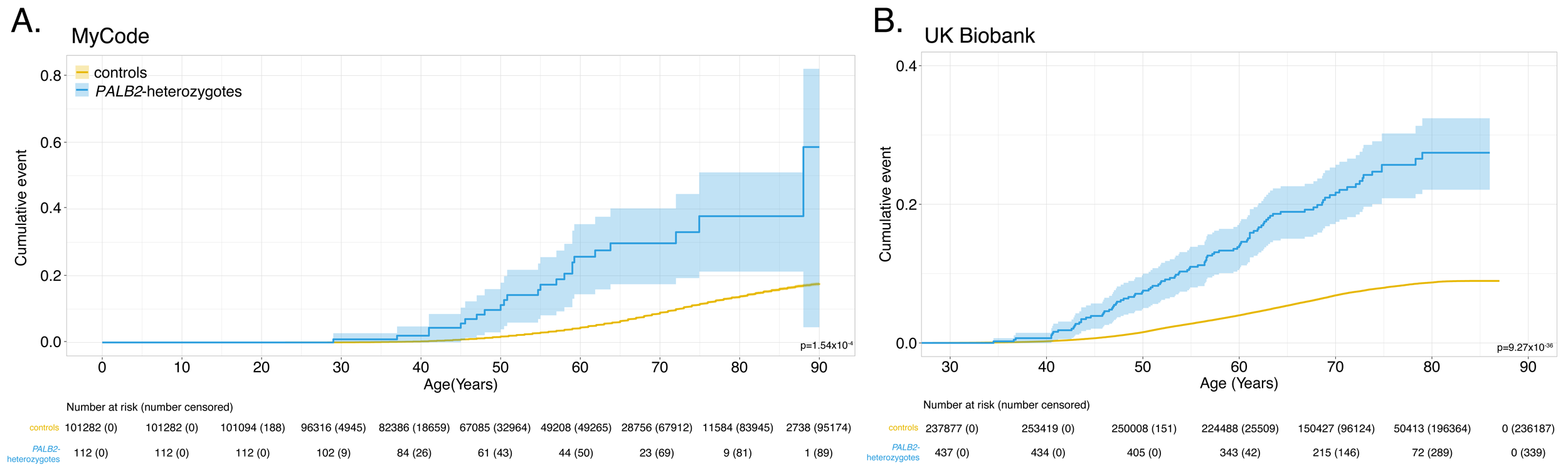
Time-to-breast-cancer in MyCode (Panel A) and UKBB (Panel B). See also **Supplemental Table 5**.

**Figure 4.**
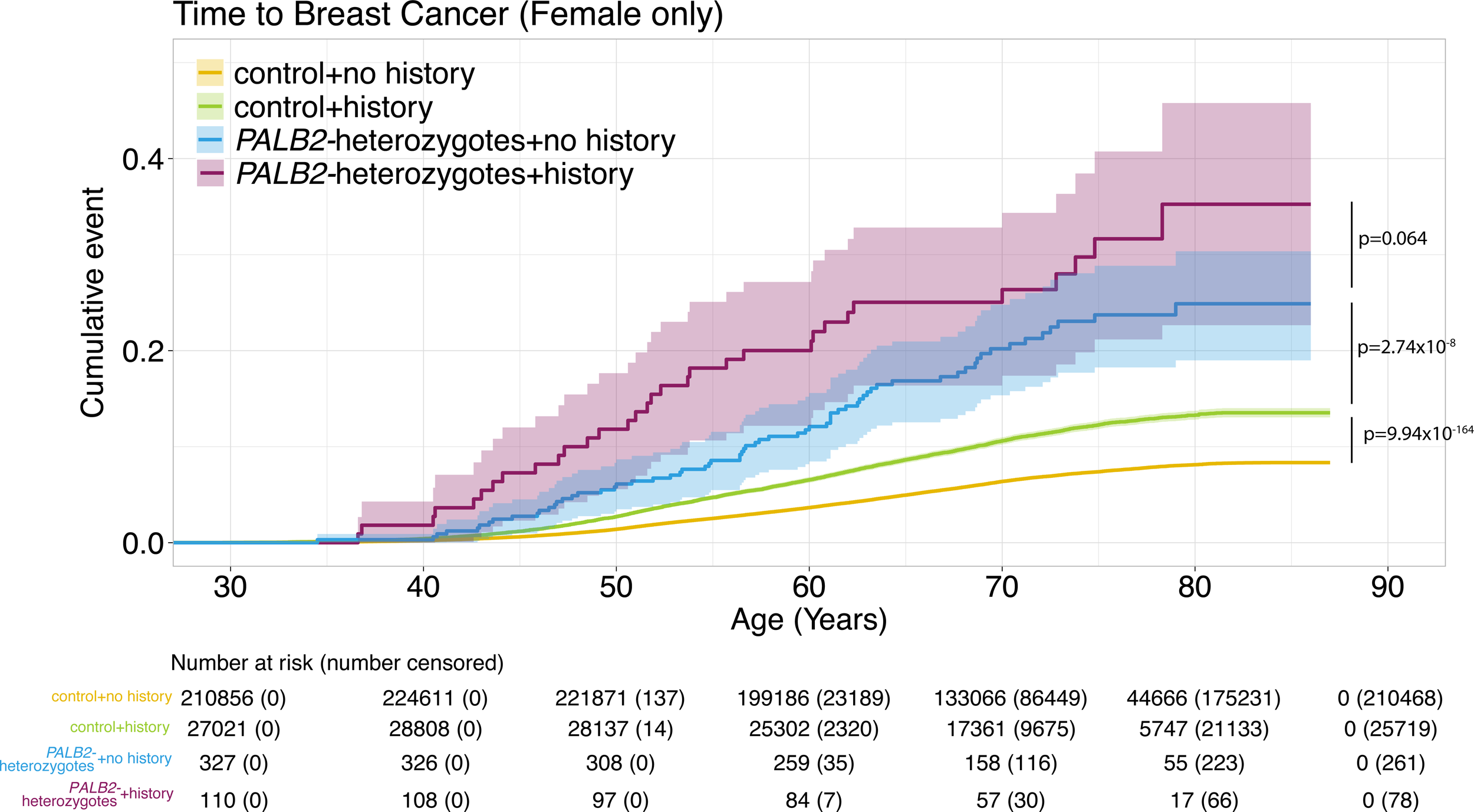
Time-to-breast-cancer (female only) in *PALB2*-heterozygotes with and without a family history of breast cancer in the UK Biobank. See also **Supplemental Table 6.**

### Time-to-pancreatic-cancer and time-to-skin cancer significantly younger in PALB2-heterozygotes vs controls in UKBB

In both cohorts, the excess odds of malignant neoplasms of digestive organs was driven primarily by pancreatic cancers (**Supplemental Figure 5)**. **Supplemental Figure 9** shows time-to-pancreatic cancer in UKBB; *PALB2*-heterozygote risk diverges significantly from controls around age 70 (P = 1.33 x 10^-6^; **Supplemental Table 7**). The excess of skin cancers was driven by both melanoma and non-melanoma cancers in UKBB; the ratio of basal/squamous keratinocyte skin cancers was similar for both *PALB2-*heterozygotes and controls in both MyCode and UKBB (2.6/1; 2.5/1; **Supplemental Data 1**).

**Supplemental Figure 10A** shows time-to-all skin cancers in UKBB for *PALB2*-heterozygotes, which was significantly younger than in controls (P = 3.64 x 10^-3^). Time-to-melanoma (**Supplemental Figure 10B**) and non-melanoma cancers (**Supplemental Figure 10C**) are also shown. **Supplemental Table 8** shows the per-decade risk of skin cancer stratified by sex and histology in UKBB.

### PALB2 p.Trp1038Ter heterozygotes have an increased risk of all cancers

**Supplemental Figure 11** displays the odds ratio of all heterozygotes, all *PALB2*-heterozygotes without *PALB2* p.Trp1038Ter and *PALB2* p.Trp1038Ter-only heterozygotes for all organ system groupings of cancer ICD codes in the UKBB. Variant-specific (*PALB2* p.Trp1038Ter-heterozygotes only) analysis shows a non-significant decreased time-to-all-cancer (**Supplemental Figure 12A; Supplemental Table 9**) and time-to-breast cancer (**Supplemental Figure 12B; Supplemental Table 10)**

### All-cause mortality was significantly increased in PALB2-heterozygotes compared to controls in both cohorts

All-cause mortality was significantly increased in *PALB2*-heterozygotes in MyCode (adjusted HR 1.67 [95% CI 1.17-2.38], P = 5.12 x 10^-3^) and UKBB (adjusted HR 1.61 [95% CI 1.33-1.94], P-value: 8.83 x 10^-7^) (**Figure 5)**. The increased mortality appeared around age 55 in MyCode and around age 65 in UKBB. **Supplemental Figure 13** shows overall mortality for all *PALB2*-heterozygotes without *PALB2* p.Trp1038Ter and *PALB2* p.Trp1038Ter-only heterozygotes vs controls. The mortality for *PALB2* p.Trp1038Ter-only heterozygotes is not significantly different to all *PALB2*-heterozygotes without *PALB2* p.Trp1038Ter, although there is a divergence around age 80 years.

**Figure 5.**
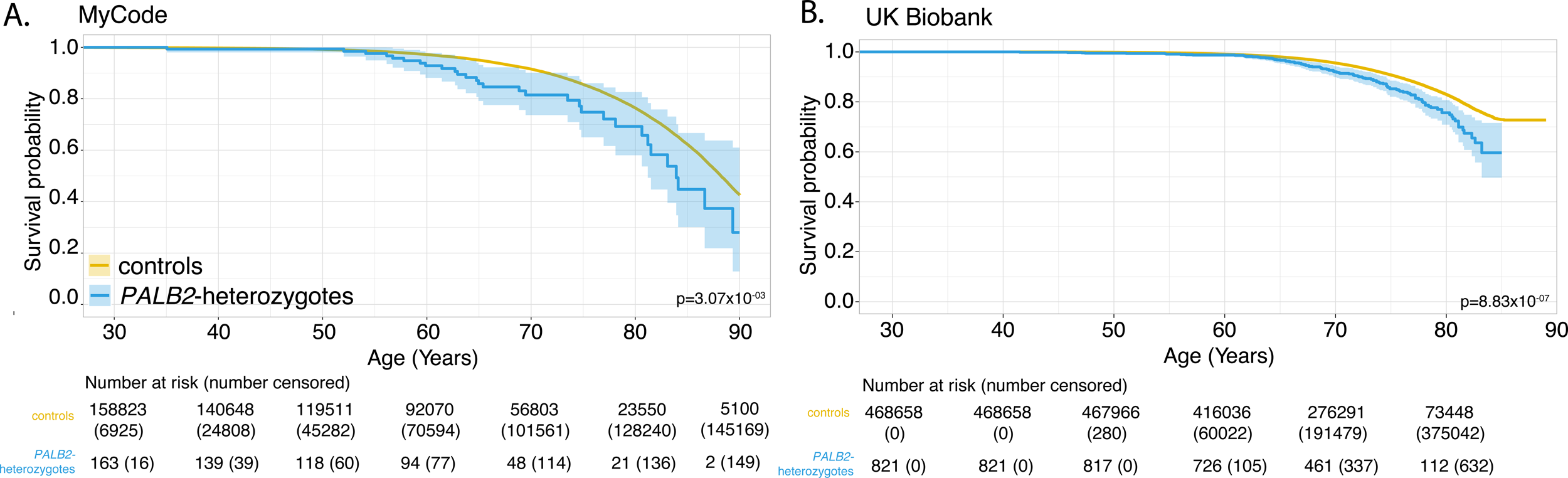
All-cause mortality in MyCode (Panel A) and in UKBB (Panel B). See also **Supplemental** Figure 13 (all-cause mortality for *PALB2* p.Trp1038Ter).

### PALB2-heterozygotes with cancer have higher mortality vs controls with cancer

There was no significant difference in overall survival for cancer-free *PALB2*-heterozygotes vs. cancer-free controls in both MyCode and UKBB. In both cohorts for all cancers, there was a significantly reduced overall survival of *PALB2*-heterozygotes with cancer vs controls with cancer (**Figure 6)**. There was decreased overall survival in *PALB2*-heterozygotes with female breast cancer vs controls with female breast cancer in UKBB but not MyCode (**Supplemental Figure 14).**

**Figure 6.**
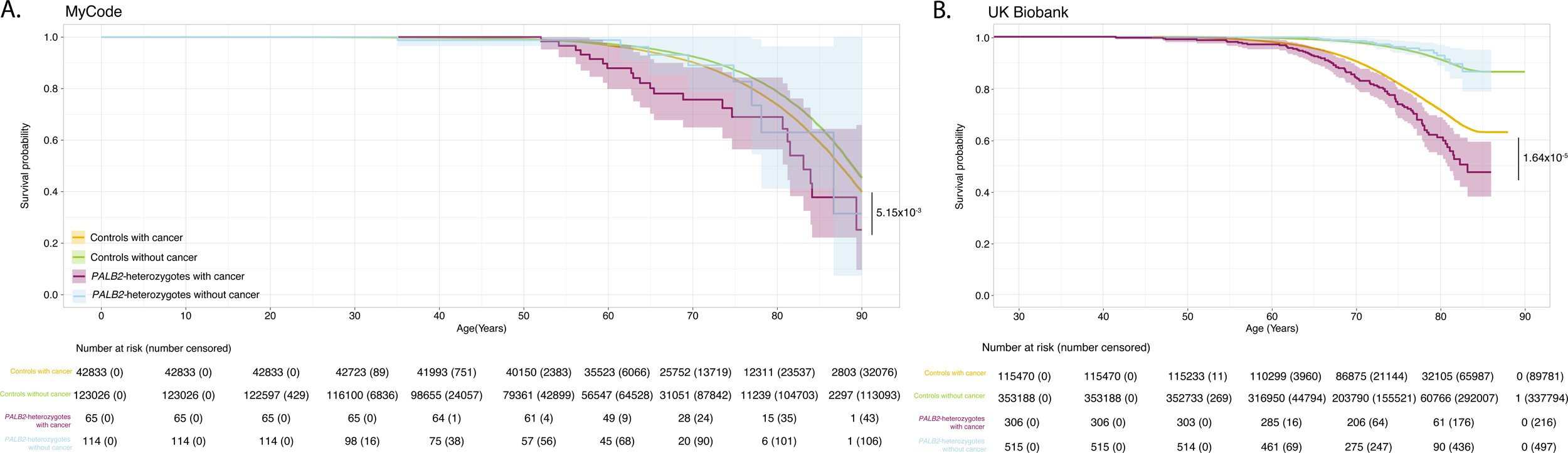
All-cause mortality for individuals with cancer in MyCode (Panel A) and in UKBB (Panel B).

## Discussion

In this case-control study, genomic ascertainment of *PALB2*-heterozygotes across two population-scale biobanks revealed an increased risk of all cancers, female breast cancer and pancreatic cancer, albeit at a lower cumulative incidence (penetrance) and odds ratio than has been previously reported from familial ascertainment. Family history of cancer increases those risks. A significant excess risk of ovarian, skin (both melanoma and non-melanoma) and neuroendocrine cancers was also observed but in one cohort only. Across all cancers, mortality in *PALB2*-heterozygotes with cancer was higher than in controls with cancer. These findings have clinical implications for individuals ascertained this way (vs. with a personal or family history of cancer).

The prevalence of *PALB2* pathogenic/likely pathogenic variants in large population databases like gnomAD is estimated at ∼0.18% (∼1/550).^18^ This is comparable to the frequency we observe in UKBB but about twice what we observed in MyCode. The difference is driven by an ∼80-fold excess of *PALB2* p.Trp1038Ter alleles in UKBB vs. MyCode, consistent with a hypothesized (but not proven) founder effect of that variant in British populations.^19^ Adequately powered analyses showed that time-to-cancer, time-to-breast cancer and mortality for *PALB2* p.Trp1038Ter was non-significantly different vs. all *PALB2*-heterozygotes (others + p.Trp1038Ter). Variant-specific (rather than just gene-specific) risk estimates should be a goal for future studies.

The risk of female breast cancer in *PALB2*-heterozygotes has been intensively studied. An international study of 524 families with a *PALB2* pathogenic variant quantified risk in women by age 80 years.^20^ Although not directly comparable, the absolute and relative risk estimates were higher than the penetrance from genomic ascertainment observed in MyCode and UKBB in this study. In contrast, a comprehensive analysis of data from the BRIDGES, CARRIERS and UKBB cohorts showed that the odds ratio from a *PALB2* pathogenic variant ascertained through phenotype (“population-type” breast cancer, independent of family history) was ∼4.3-fold,^21^ roughly comparable to the odds ratio in this investigation. We also observed excess risk of DCIS in both cohorts, confirming previous reports.^22,23^ Family history increases risk of cancer, including those with already high risk, such as *PALB2*-heterozygotes. These risks are very high (∼80% by age 80 years) in phenotypically ascertained cohorts^24^ and lower when genotypically ascertained (∼27-38% by age 80 years; **Figure 3**). Risk data on other cancer types are more uncertain. Our estimates of pancreatic and ovarian cancer risk are roughly comparable to those of Yang et al,^20^ but are constrained by the limited number of observations. There is a known healthy volunteer effect in UKBB that may explain the much lower risk of all cancers and breast cancer by age 80 (**Figure 2**; **Figure 3**).^25^ In line with our observations, there is no consistent evidence that *PALB2*-heterozygotes have an increased risk of colorectal or prostate cancers,^20,26^ comparable to our observations. The quantification of risk from all (aggregated) cancer types to our knowledge has not been previously reported.

Cancer studies using genomic ascertainment are not biased by phenotype and thus can uncover novel or under-recognized tumor associations. Among cancers in organ systems with at least three observations, there was excess risk in *PALB2*-heterozygotes for ovarian cancer, liver and intrahepatic bile duct cancers, neuroendocrine cancers, melanoma and non-melanoma skin cancers. However, this should be tempered by the observation in only one cohort and/or lack of significance after Bonferroni correction (∼3.6 x 10^-3^ = 0.05/14, with 14 organ systems tested). The excess skin cancer is observed in the UKBB cohort; there was also increased risk for benign and in situ skin carcinomas in UKBB. Although germline *PALB2* pathogenic variants have been reported in patients with uveal melanoma,^27^ sequencing studies of phenotypically ascertained cohorts of cutaneous melanoma have not demonstrated increased risk in *PALB2*-heterozygotes.^28,29^ In addition, 94% (n=17) of the melanomas were a participant’s first diagnosed cancer, suggesting that surveillance bias was unlikely. One melanoma was classified as lentigo maligna, a subtype associated with high cumulative solar damage and not typically linked to cancer predisposition syndromes.^30^ In contrast, most melanomas observed in the *PALB2*-heterozygote group were non-lentigo maligna subtypes, which are more often associated with low cumulative solar damage and, in some cases, hereditary cancer predisposition **(Supplemental Data 1**).^30^ We are not aware of studies investigating *PALB2* risk of non-melanoma skin cancer or in benign skin neoplasms. Malignant neuroendocrine tumors of various histologies have been reported in *PALB2*-heterozygotes^31–34^ but we are not aware of any literature reporting *PALB2*-associated benign neuroendocrine tumors; to our knowledge, large-scale studies like ours have not been performed on either tumor type. Replication in additional cohorts and/or evidence of a Knudson second hit would help establish these tumor types as *PALB2*-associated.

We observed significantly worse overall survival for *PALB2*-heterozygotes in both cohorts, driven by cancer mortality. There are conflicting data on mortality of *PALB2*-associated cancers. Studies of breast^35–38^ cancer have shown more aggressive disease and worse survival in *PALB2-*heterozygotes. In contrast, a large population-based study of genetically tested breast, colorectal and pancreatic cancers found similar mortality in *PALB2-*heterozygotes vs controls.^39^ The difference in cancer mortality may be explained by ascertainment. In the current investigation, individuals were ascertained based on *PALB2* genotype, regardless of cancer status; presumably, there were numerous individuals who underwent cancer chemotherapy without the knowledge of their *PALB2*-genotype. In the work of Veenstra et al, individuals were ascertained based on cancer diagnosis (through the SEER registry from 2013-2019) and previous genetic testing status; presumably many individuals had therapeutic decisions (e.g., FDA-approval of PARP-inhibitors December 2014 for ovarian cancer and January 2018 for breast cancer) based on their genotype and may have benefitted from it.

### Limitations

There are limitations to these retrospective analyses. Absolute risk was not quantified. UKBB and MyCode are predominantly of European ancestry. Copy-number variants in *PALB2* were not evaluated due to limited data availability in UKBB. Enrollment may have been biased since individuals with conditions leading to death or disabilities would be less likely to participate. A healthy volunteer bias has been documented in the UKBB.^25^

### Conclusions

Genomic ascertainment of *PALB2*-heterozygotes shows that breast cancer risk in individuals is comparable to risk from phenotypic ascertainment but lower than that observed with familial ascertainment. All-cause mortality was significantly increased in *PALB2*-heterozygotes compared to controls in both cohorts, driven by cancer mortality. These findings underscore the importance of ongoing surveillance and informed management of *PALB2*-heterozygote individuals identified through genomic screening.

The content of this publication does not necessarily reflect the views or policies of the Department of Health and Human Services, nor does mention of trade names, commercial products or organizations imply endorsement by the U.S. Government.

This research was supported, in part, by the Intramural Research Program of the National Institutes of Health (NIH). The contributions of the NIH authors were made as part of their official duties as NIH federal employees, are in compliance with agency policy requirements, and are considered Works of the United States Government. However, the findings and conclusions presented in this paper are those of the authors and do not necessarily reflect the views of the NIH or the U.S. Department of Health and Human Services.

The authors would like to acknowledge the participants of the MyCode Community Initiative for the use of their health and genomic information, without whom this study would not be possible. The patient enrollment and exome sequencing were funded by the Regeneron Genetics Center. Data for this project was made possible by the Geisinger-Regeneron DiscovEHR Collaboration.

## Data Availability

UK Biobank data are publicly available through UK Biobank.
Geisinger MyCode data to reproduce the results are available to qualified academic non-commercial researchers under a data access
agreement.

## Acknowledgments

This work was supported by the Intramural Research Program of the Division of Cancer Epidemiology and Genetics of the National Cancer Institute, Bethesda, MD and utilized the computational resources of the NIH High-Performance Computing Biowulf cluster. This research has been conducted using the UK Biobank Resources under application 54389. MT was supported by the NIHR Cambridge Biomedical Research Centre (NIHR203312)

## Key objective

To quantify cancer incidence, mortality risk, and age-specific penetrance associated with heterozygous pathogenic or likely pathogenic *PALB2* variants identified through genomic ascertainment, and to assess modification of these risks by family history of cancer.

## Knowledge generated

Across two large population-based cohorts, *PALB2*-heterozygotes had significantly increased odds of any cancer, breast cancer, and pancreatic cancer, and higher age-specific hazard of cancer and higher all-cause mortality compared with non-carriers. Family history further elevated cancer risk. Age-specific penetrance estimates were lower than those reported from familial ascertainment.

## Supplemental Figures

**Supplemental Figure 1.** Power as a function of risk (odds ratio) in MyCode for a range of cancer rates. Prevalence data from cohort-specific frequency of *PALB2*-heterozygotes (**Table 1**). Dark gray line represents 80% power, and light gray line represents 90% power.

**Supplemental Figure 2.** Power as a function of risk (odds ratio) in UK Biobank for a range of cancer rates. Prevalence data from cohort-specific frequency of *PALB2*-heterozygotes (**Table 1**). Dark gray line represents 80% power, and light gray line represents 90% power.

**Supplemental Figure 3.** Power as a function of risk (odds ratio) in UK Biobank for a range of cancer rates. Prevalence data from cohort-specific frequency of *PALB2* p.Trp1038Ter. Dark gray line represents 80% power, and light gray line represents 90% power.

**Supplemental Figure 4**. Power as a function of risk (odds ratio) in UK Biobank for a range of cancer rates. Prevalence data from cohort-specific frequency of *PALB2*-heterozygotes without *PALB2* p.Trp1038Ter. Dark gray line represents 80% power, and light gray line represents 90% power.

**Supplemental Figure 5.** Expansion of odds ratio for *PALB2*-heterozygotes for organ system groupings of cancer ICD codes with a significant excess of risk in MyCode and UK Biobank. CI: 95% confidence interval; OR: odds ratio. All organs are listed for each organ system, even if no tumors were reported (*e.g.,* C58: malignant neoplasm of placenta)

**Supplemental Figure 6.** Odds ratio of *PALB2*-heterozygotes for carcinoma *in situ* codes in MyCode and UK Biobank

**Supplemental Figure 7**. Odds ratio of *PALB2*-heterozygotes for benign tumor codes in MyCode and UK Biobank

**Supplemental Figure 8.** Odds ratio of *PALB2*-heterozygotes for tumors of uncertain behavior in MyCode and UK Biobank

**Supplemental Figure 9.** Time-to-pancreatic-cancer in *PALB2*-heterozygotes in the UK Biobank. See also **Supplemental Table 7**

**Supplemental Figure 10.** Time-to-skin-cancer (panel A), time-to-melanoma (panel B) and time-to-non-melanoma (Panel C) in *PALB2*-heterozygotes in the UK Biobank. See also **Supplemental Table 8.**

**Supplemental Figure 11.** Odds ratio for all *PALB2*-heterozygotes, all *PALB2*-heterozygotes without Trp1038Ter and Trp1038Ter-only heterozygotes for organ system groupings of cancer ICD codes with a significant excess of risk in UK Biobank. CI: 95% confidence interval; OR: odds ratio

**Supplemental Figure 12.** Time-to-cancer for All *PALB2*-heterozygotes without *PALB2* Trp1038Ter and *PALB2* Trp1038Ter heterozygotes vs controls (panel A) and time-to-breast cancer (female only) for All *PALB2*-heterozygotes without *PALB2* Trp1038Ter and *PALB2* Trp1038Ter heterozygotes vs controls (panel B). See also **Supplemental Table 9** and **Supplemental Table 10.**

**Supplemental Figure 13.** All-cause mortality in UK Biobank for All *PALB2*-heterozygotes without *PALB2* Trp1038Ter and *PALB2* Trp1038Ter heterozygotes vs controls. See also **Figure 5** (all-cause mortality for *PALB2*-heterozygotes).

**Supplemental Figure 14.** All-cause mortality in *PALB2*-heterozygotes with and without breast cancer vs controls with and without breast cancer in MyCode (panel A) and UK Biobank (panel B).

## Supplemental Tables

**Supplemental Table 1.** Allele counts and frequencies of *PALB2* pathogenic (P) and likely pathogenic (LP) variants in UK Biobank and Geisinger MyCode. No *PALB2* p.Trp1038Ter variants were observed in the MyCode cohort.

**Supplemental Table 2.** List of all *PALB2* pathogenic and likely pathogenic variants found in the study

**Supplemental Table 3.** Demographics of *PALB2*-heterozygotes vs controls. LP: likely pathogenic variant; P: pathogenic variant

**Supplemental Table 4.** Per-decade risk of all cancers stratified by sex in MyCode and UK Biobank

**Supplemental Table 5.** Per-decade risk of breast cancer in MyCode and UK Biobank

**Supplemental Table 6.** Per-decade risk of female breast cancer in women with and without a family history of breast cancer in UK Biobank

**Supplemental Table 7** Per-decade risk of pancreatic cancer in UK Biobank

**Supplemental Table 8** Per-decade risk of skin cancer stratified by sex and histology in UK Biobank

**Supplemental Table 9.** Per-decade risk of all cancers in UK Biobank for *PALB2* Trp1038Ter heterozygotes

**Supplemental Table 10.** Per-decade risk of all breast cancer (female only) in UK Biobank for *PALB2* Trp1038Ter heterozygotes

## Supplemental Data

**Supplemental Data 1.** Histology and age of diagnosis of *PALB2*-associated melanoma in the UK Biobank.

