## Supplemental Data 1 for "Genomic ascertainment of *PALB2*-related cancer predisposition"

**Supplemental Data 1:** Histology and age of diagnosis of *PALB2*-associated melanoma in the UK Biobank. Frequency of Non-melanoma skin cancer and uveal melanoma in the UK Biobank.

**UK Biobank**

| ICD10 | age_at_diagnosis | histology | Cohort |
| --- | --- | --- | --- |
| C43.7 Malignant melanoma of lower limb, including hip | 54 | Acral lentiginous melanoma, malig. | UKBB |
| C43.6 Malignant melanoma of upper limb, including shoulder | 58 | Lentigo maligna melanoma | UKBB |
| C43.7 Malignant melanoma of lower limb, including hip | 39 | Malignant melanoma, NOS | UKBB |
| C43.6 Malignant melanoma of upper limb, including shoulder | 63 | Malignant melanoma, NOS | UKBB |
| C43.3 Malignant melanoma of other and unspecified parts of face | 60 | Malignant melanoma, NOS | UKBB |
| C43.3 Malignant melanoma of other and unspecified parts of face | 65 | Malignant melanoma, NOS | UKBB |
| C43.5 Malignant melanoma of trunk | 60 | Malignant melanoma, NOS | UKBB |
| C43.6 Malignant melanoma of upper limb, including shoulder | 72 | Malignant melanoma, NOS | UKBB |
| C43.9 Malignant melanoma of skin, unspecified | 66 | Malignant melanoma, NOS | UKBB |
| C43.5 Malignant melanoma of trunk | 53 | Nodular melanoma | UKBB |
| C43.7 Malignant melanoma of lower limb, including hip | 41 | Nodular melanoma | UKBB |
| C69.3 Choroid | 65 | Spindle cell melanoma, NOS | UKBB |
| C43.6 Malignant melanoma of upper limb, including shoulder | 63 | Superficial spreading melanoma | UKBB |
| C43.5 Malignant melanoma of trunk | 53 | Superficial spreading melanoma | UKBB |
| C43.5 Malignant melanoma of trunk | 74 | Superficial spreading melanoma | UKBB |
| C43.7 Malignant melanoma of lower limb, including hip | 31 | Superficial spreading melanoma | UKBB |
| C43.3 Malignant melanoma of other and unspecified parts of face | 58 | Superficial spreading melanoma | UKBB |
| 1725 Malignant melanoma of trunk, except scrotum | 48 | Superficial spreading melanoma | UKBB |

**Non-melanoma skin cancer**

*Basal cell carcinoma* (histology codes 8090)

Controls: 29269 (6.2%)

*PALB2*: 68 (8.3%)

*Squamous cell carcinoma* (histology codes 8070)

Controls: 11259 (2.4%)

*PALB2*: 27 (3.3%)

**Uveal melanoma (C69.3)**

There is total 206 individuals in controls with uveal melanoma and 1 in *PALB2*

That 1 individual with uveal melanoma does not have *BAP1* P/LP variants
