## Supplemental Figure 14 for "Genomic ascertainment of *PALB2*-related cancer predisposition"

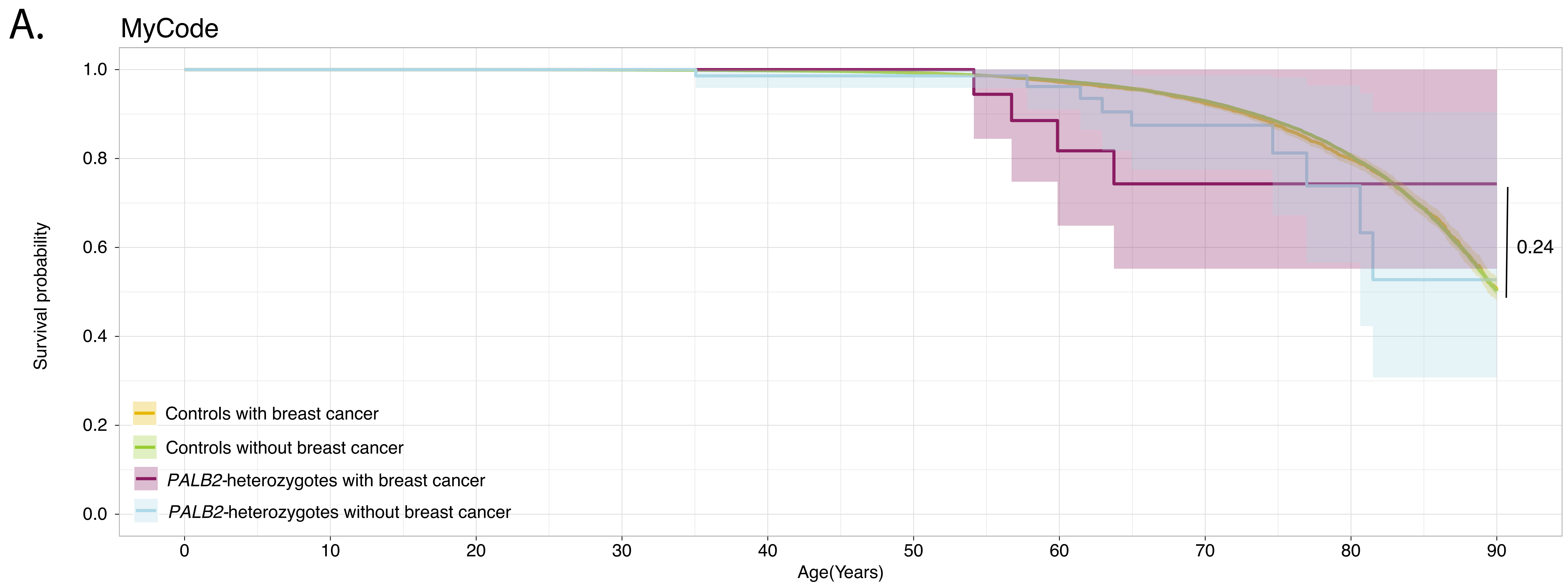


Number at risk (number censored)

| | 0 | 10 | 20 | 30 | 40 | 50 | 60 | 70 | 80 | 90 |
|---|---|---|---|---|---|---|---|---|---|---|
| Controls with breast cancer | 6146 (0) | 6146 (0) | 6146 (0) | 6141 (3) | 6087 (50) | 5833 (271) | 5024 (966) | 3530 (2248) | 1641 (3783) | 394 (5049) |
| Controls without breast cancer | 95136 (0) | 95136 (0) | 94948 (188) | 90198 (4885) | 76489 (18448) | 62190 (32430) | 45886 (47807) | 27243 (64758) | 11204 (78341) | 2695 (86902) |
| *PALB2*-heterozygotes with breast cancer | 23 (0) | 23 (0) | 23 (0) | 23 (0) | 22 (1) | 20 (3) | 12 (8) | 6 (13) | 3 (16) | 1 (19) |
| *PALB2*-heterozygotes without breast cancer | 89 (0) | 89 (0) | 89 (0) | 80 (9) | 63 (25) | 46 (42) | 39 (48) | 20 (64) | 8 (74) | 1 (80) |

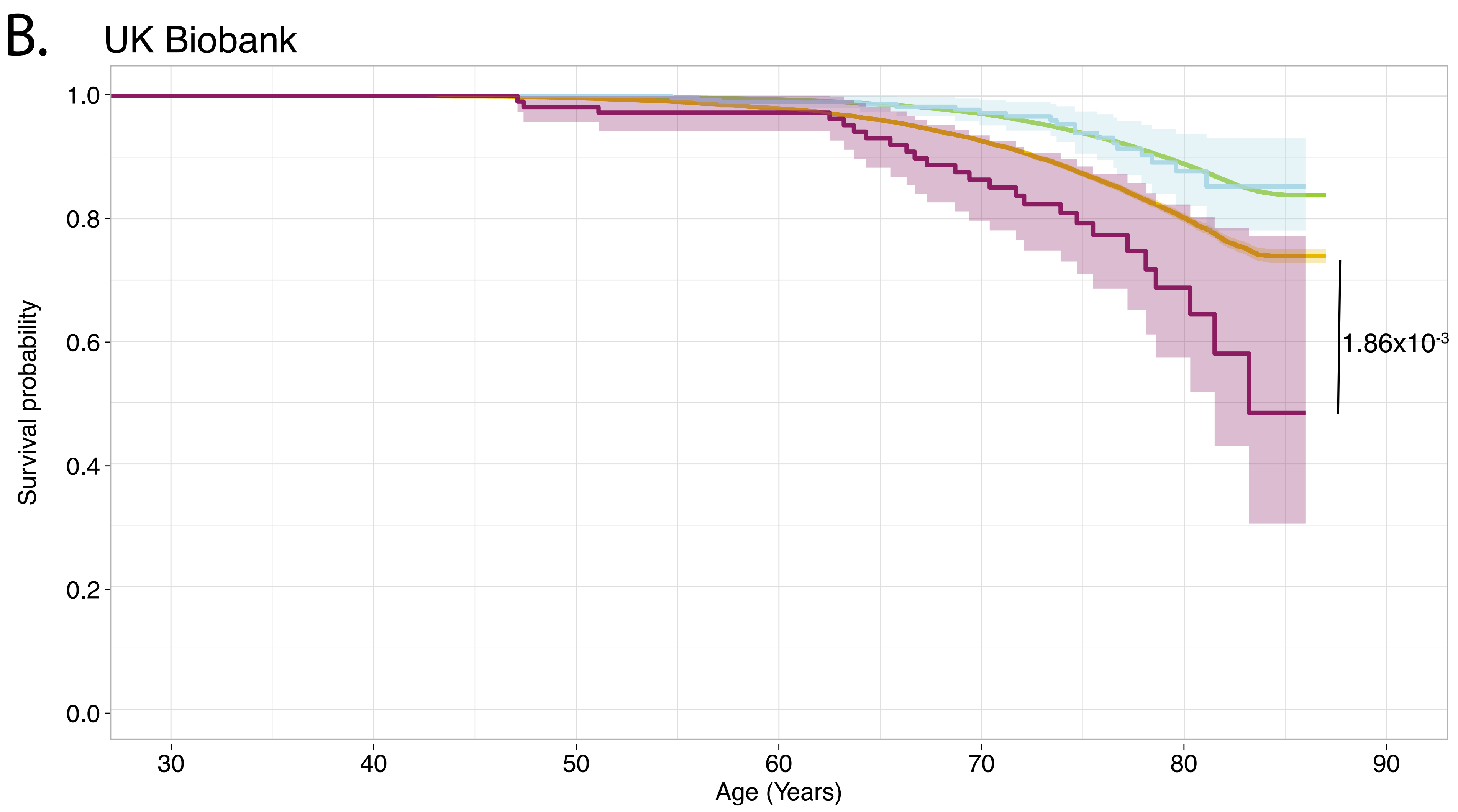


Number at risk (number censored)

| | 30 | 40 | 50 | 60 | 70 | 80 | 90 |
|---|---|---|---|---|---|---|---|
| Controls with breast cancer | 20020 (0) | 20020 (0) | 19958 (3) | 18820 (1077) | 14078 (5172) | 4821 (13625) | 0 (17213) |
| Controls without breast cancer | 233996 (0) | 233996 (0) | 233721 (151) | 213482 (25033) | 143172 (93291) | 44464 (186266) | 0 (219992) |
| *PALB2*-heterozygotes with breast cancer | 111 (0) | 111 (0) | 109 (0) | 103 (9) | 71 (31) | 19 (73) | 0 (86) |
| *PALB2*-heterozygotes without breast cancer | 326 (0) | 326 (0) | 326 (0) | 289 (42) | 188 (138) | 62 (265) | 0 (307) |
