## Supplementary figures and images for "Genomic ascertainment of *PALB2*-related cancer predisposition"

### Supplemental Figure 1

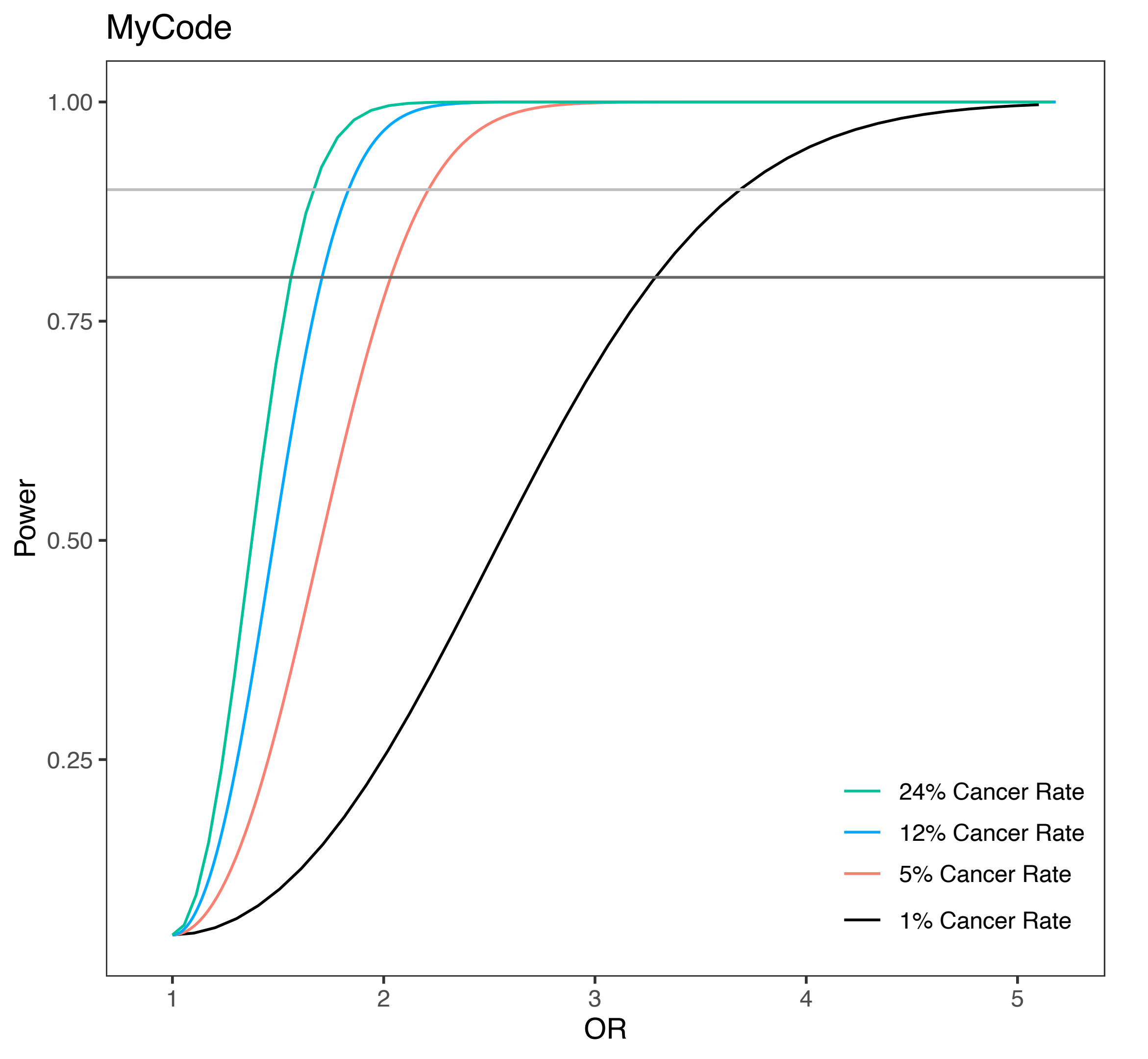

### Supplemental Figure 2

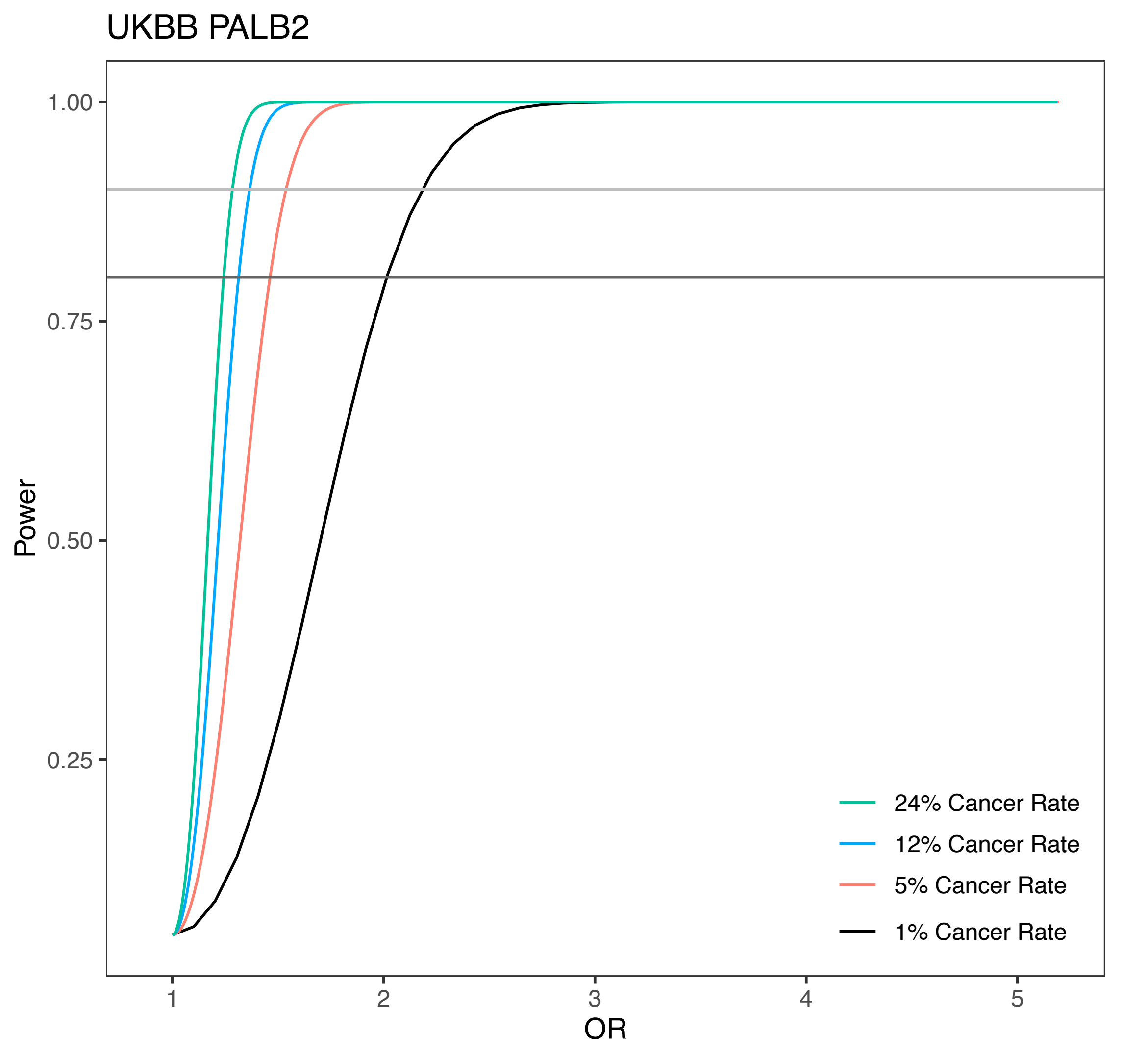

### Supplemental Figure 3

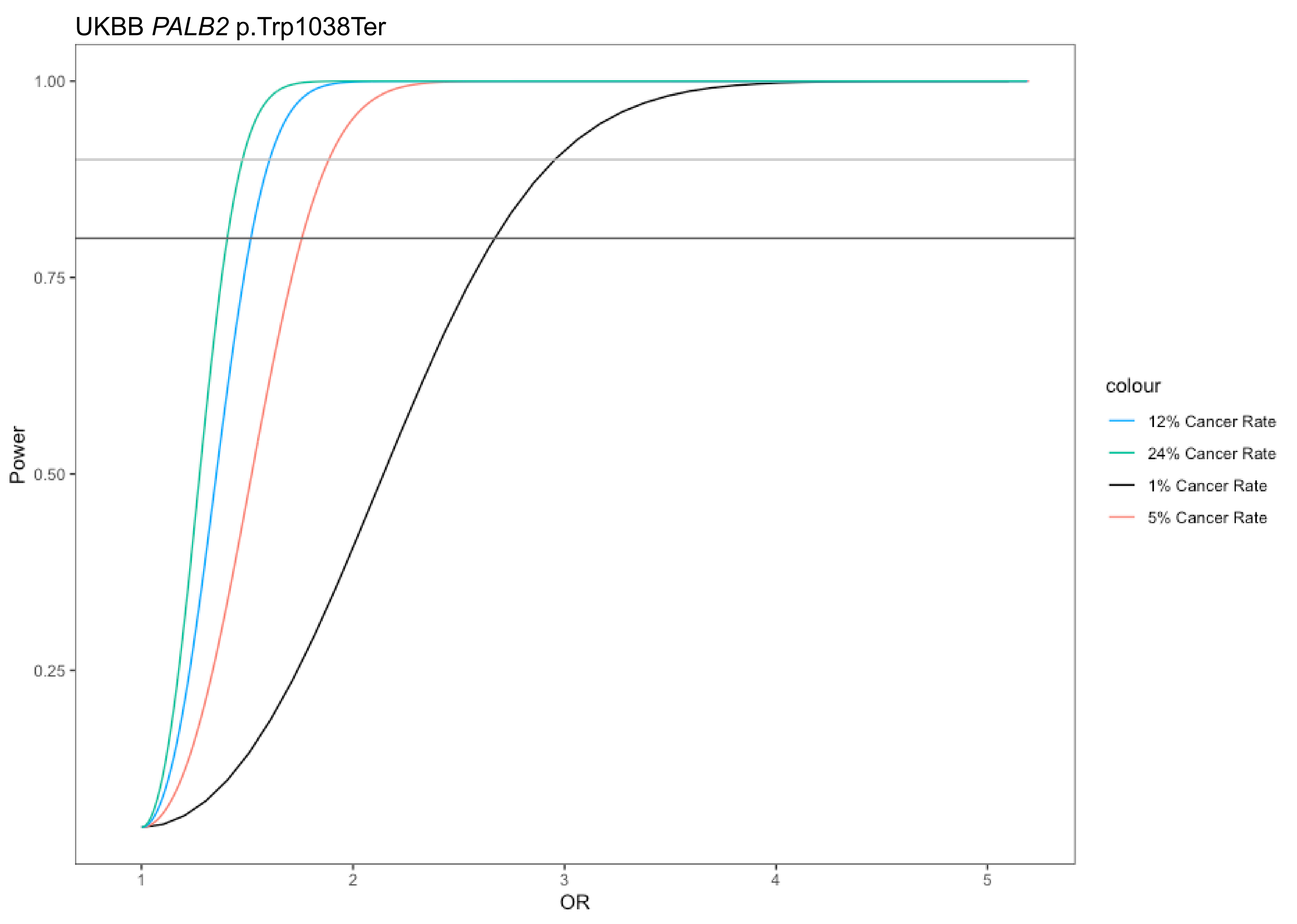

### Supplemental Figure 4

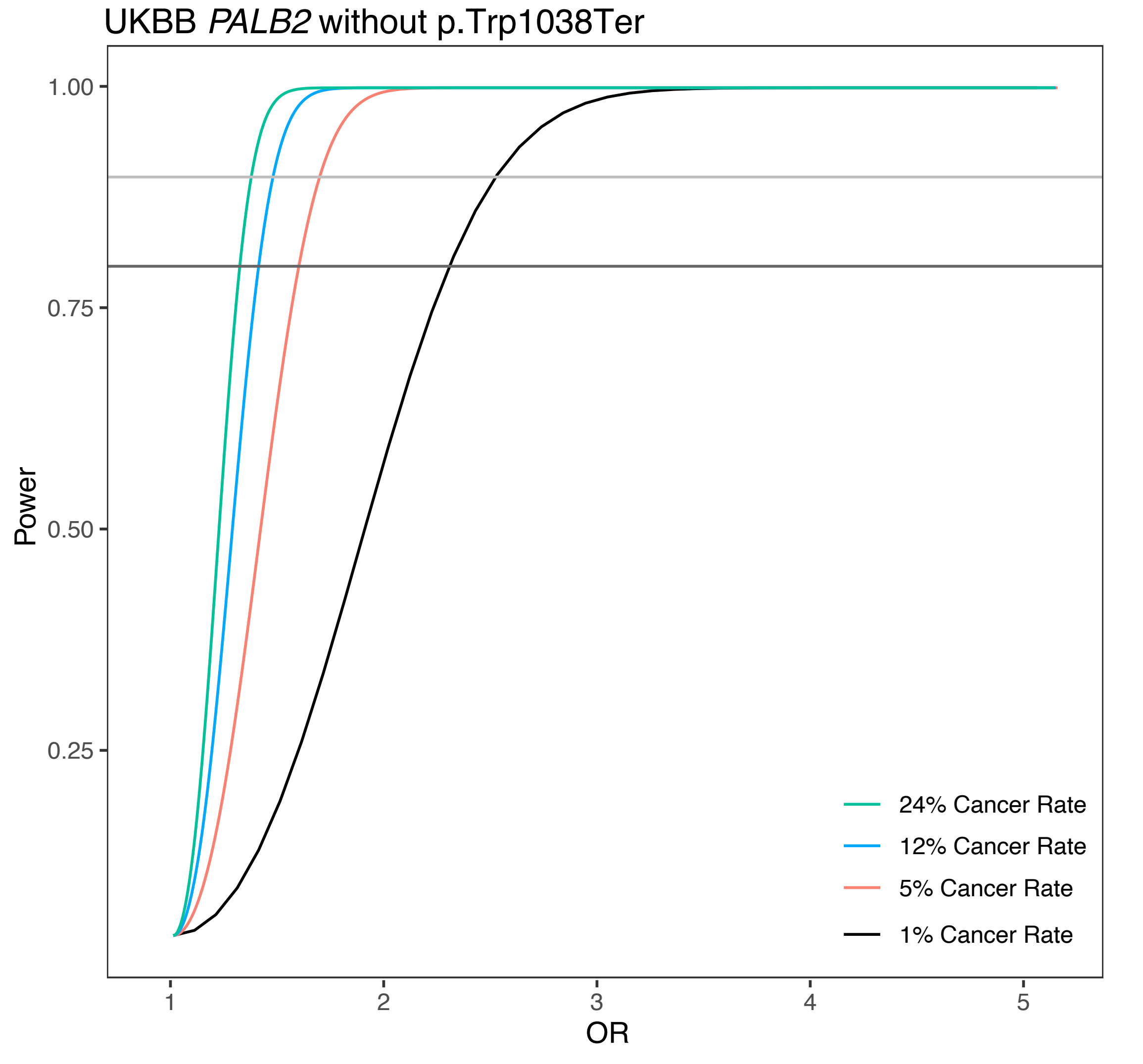

### Supplemental Figure 8

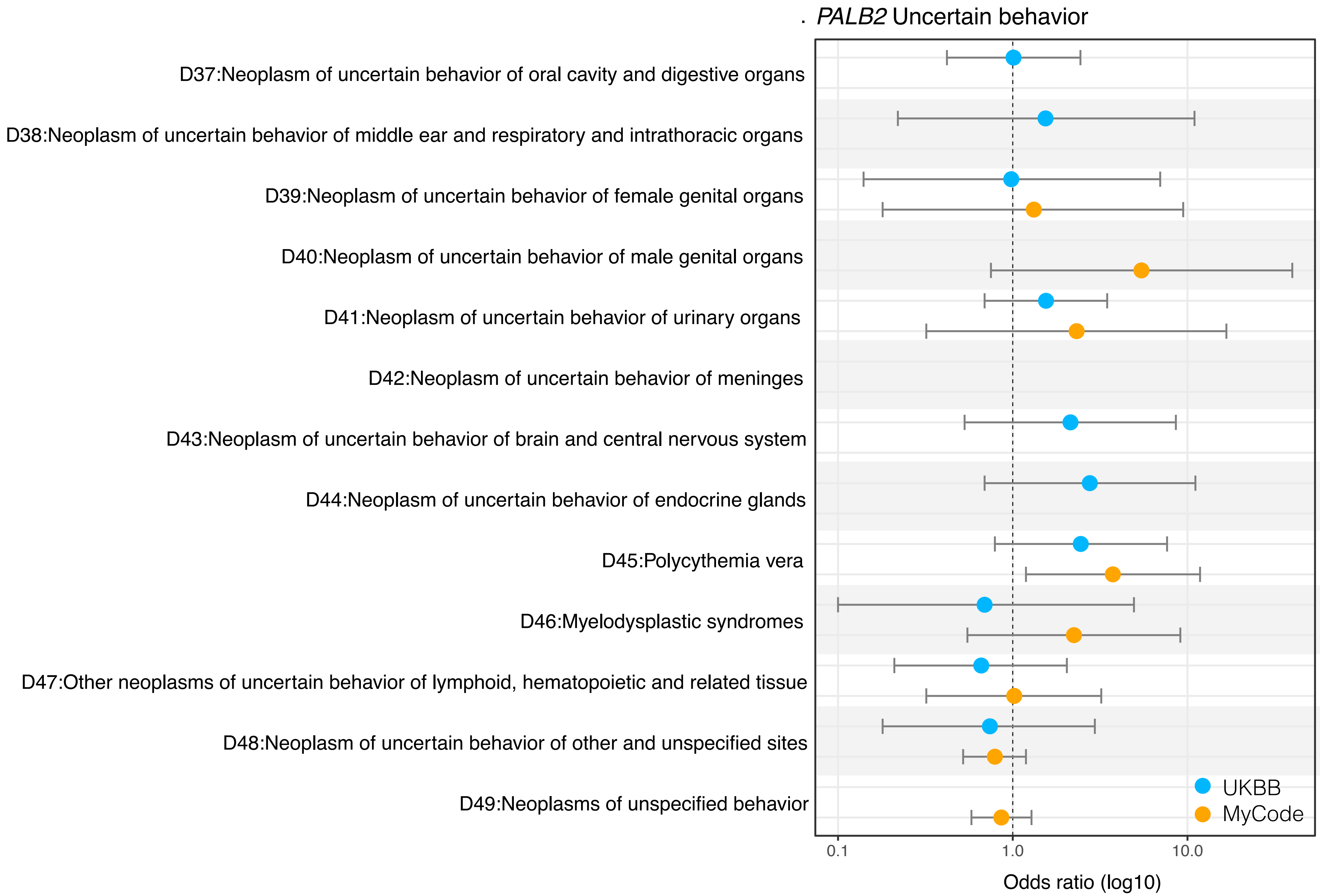


| | controls(%) | heterozygotes(%) | OR [95% CI] | p−value |
|---|---|---|---|---|
| D37 | 2866(0.61) | 5(0.61) | 1.01[0.42–2.44] | 0.98 |
| | 930(0.56) | 0(0) | NA | NA |
| D38 | 387(0.08) | 1(0.12) | 1.54[0.22–10.97] | 0.67 |
| | 265(0.159) | 0(0) | NA | NA |
| D39 | 600(0.23) | 1(0.23) | 0.98[0.14–6.98] | 0.98 |
| | 725(0.715) | 1(0.89) | 1.32[0.18–9.45] | 0.78 |
| D40 | 203(0.09) | 0(0) | NA | NA |
| | 180(0.278) | 1(1.49) | 5.45[0.75–39.8] | 0.09 |
| D41 | 2242(0.48) | 6(0.73) | 1.55[0.69–3.47] | 0.29 |
| | 446(0.268) | 1(0.56) | 2.32[0.32–16.7] | 0.40 |
| D42 | 104(0.02) | 0(0) | NA | NA |
| | 83(0.05) | 0(0) | NA | NA |
| D43 | 540(0.12) | 2(0.2) | 2.14[0.53–8.58] | 0.28 |
| | 103(0.062) | 0(0) | NA | NA |
| D44 | 417(0.09) | 2(0.24) | 2.76[0.69–11.1] | 0.15 |
| | 498(0.3) | 0(0) | NA | NA |
| D45 | 704(0.15) | 3(0.37) | 2.45[0.79–7.64] | 0.12 |
| | 821(0.49) | 3(1.68) | 3.74[1.19–11.8] | 0.02 |
| D46 | 845(0.18) | 1(0.12) | 0.69[0.1–4.94] | 0.72 |
| | 946(0.57) | 2(1.117) | 2.24[0.55–9.1] | 0.26 |
| D47 | 2659(0.57) | 4(0.49) | 0.66[0.21–2.04] | 0.47 |
| | 3039(1.832) | 3(1.68) | 1.02[0.32–3.21] | 0.98 |
| D48 | 1575(0.34) | 2(0.24) | 0.74[0.18–2.95] | 0.88 |
| | 34244(20.6) | 29(16.2) | 0.79[0.52–1.19] | 0.26 |
| D49 | 0(0) | 0(0) | NA | NA |
| | 32861(19.81) | 30(16.75) | 0.86[0.58–1.28] | 0.46 |

### Supplemental Figure 9

Time to Pancreatic Cancer

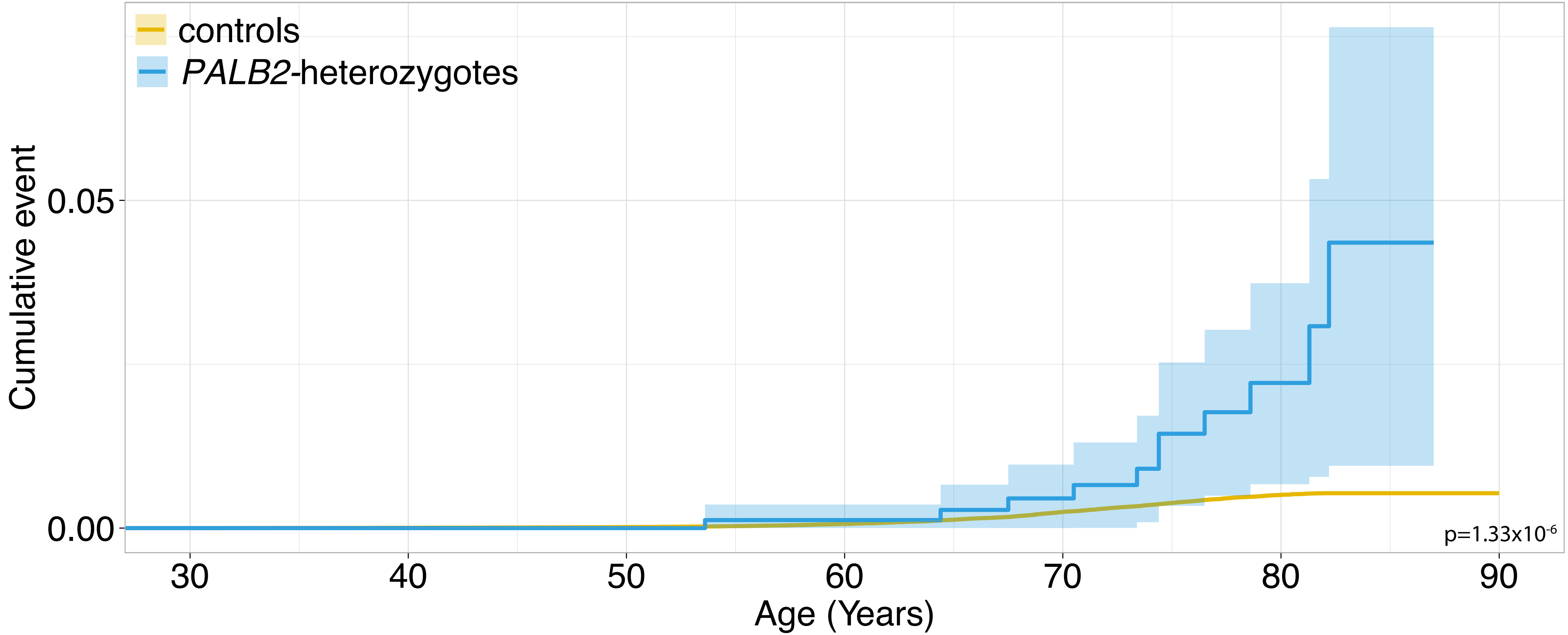


Number at risk (number censored)

| | | | | | | | |
|---|---|---|---|---|---|---|---|
| controls | 463034 (0) | 468638 (0) | 468354 (280) | 430254 (49593) | 300692 (182215) | 108409 (381084) | 1 (467165) |
| *PALB2*-heterozygotes | 821 (0) | 821 (0) | 821 (0) | 750 (90) | 513 (330) | 188 (671) | 0 (810) |

### Supplemental Figure 10

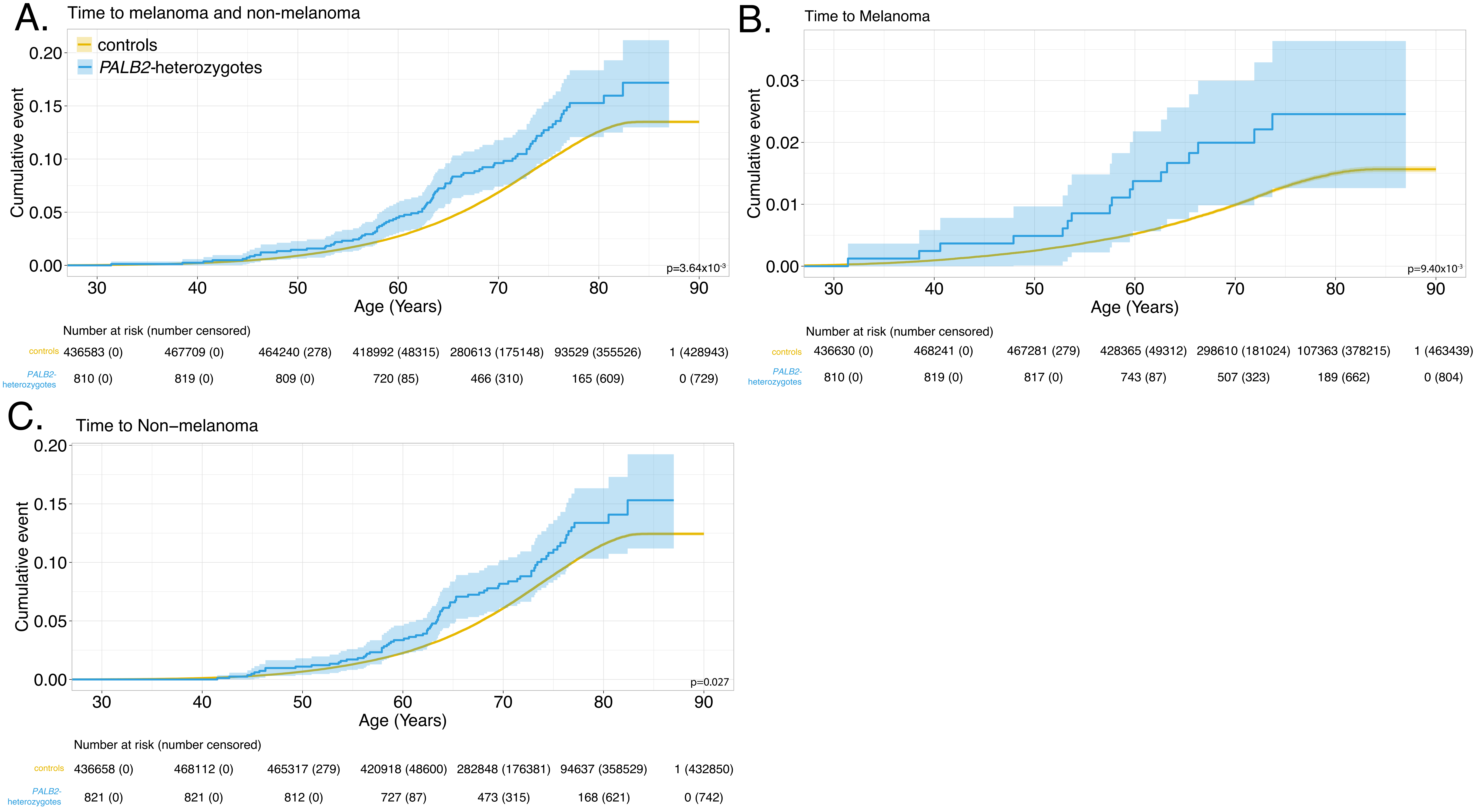

### Supplemental Figure 13

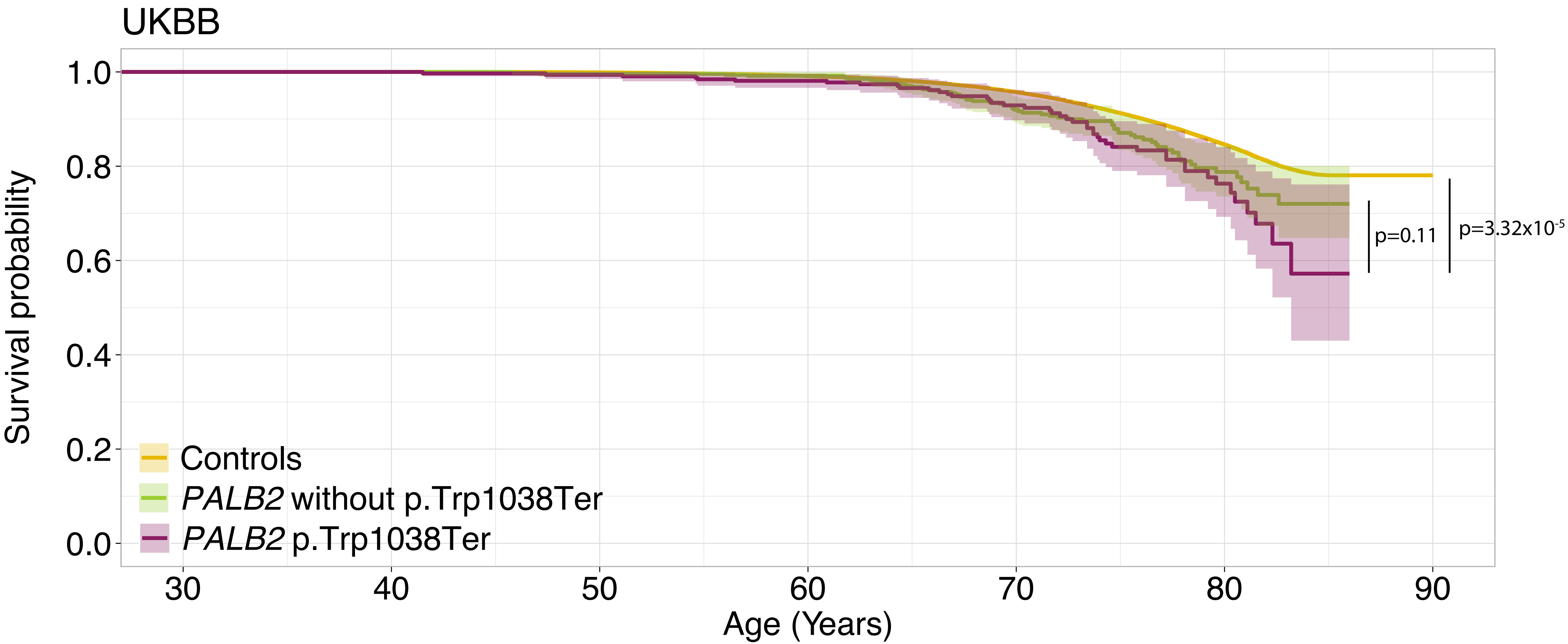


Number at risk (number censored)

| | | | | | | | |
|---|---|---|---|---|---|---|---|
| Controls | 468658 (0) | 468658 (0) | 467966 (280) | 427249 (48754) | 290665 (176665) | 92871 (357994) | 1 (427575) |
| *PALB2* het without p.Trp1038Ter | 502 (0) | 502 (0) | 500 (0) | 457 (54) | 300 (185) | 94 (372) | 0 (439) |
| *PALB2* p.Trp1038Ter | 319 (0) | 319 (0) | 317 (0) | 289 (31) | 181 (126) | 57 (240) | 0 (274) |
