## Supplemental Figure 12 for "Genomic ascertainment of *PALB2*-related cancer predisposition"

A.

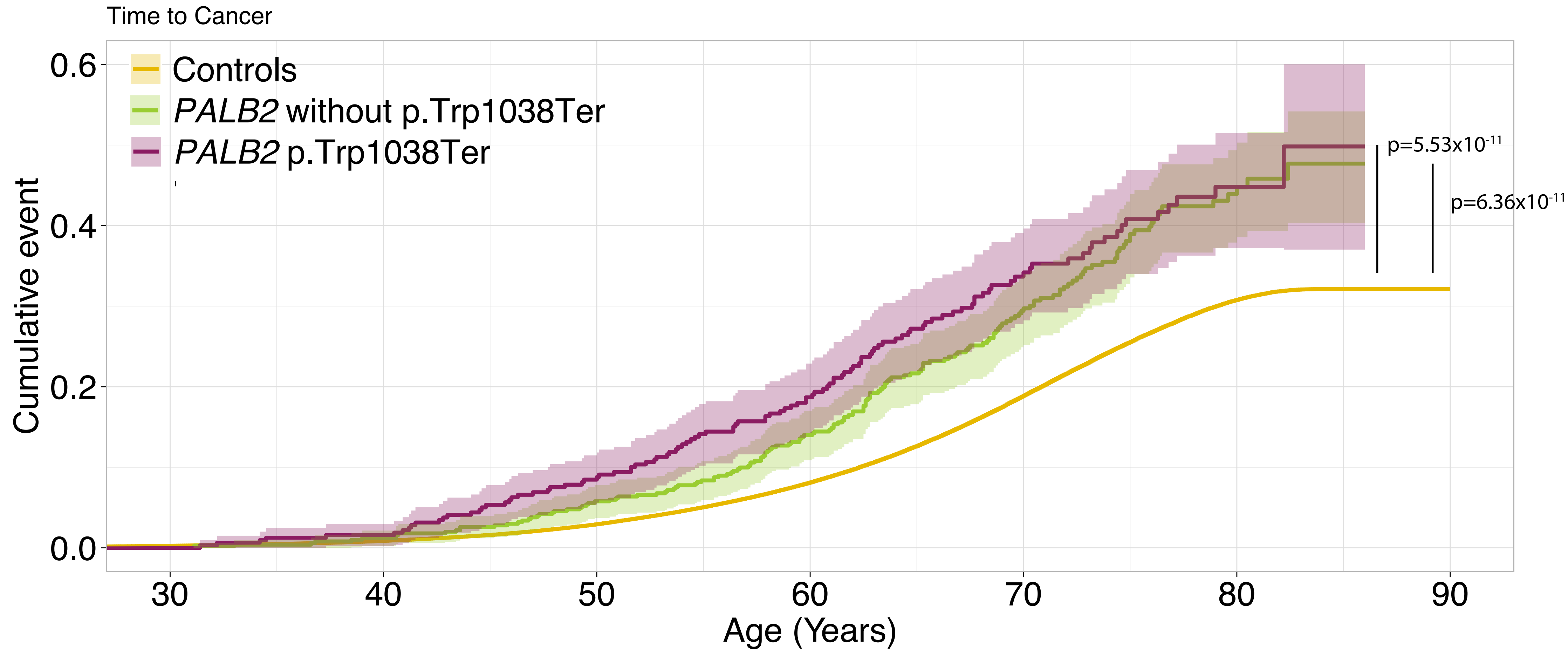


Number at risk (number censored)

| | | | | | | | |
|---|---|---|---|---|---|---|---|
| Controls | 435908 (0) | 465000 (0) | 455205 (272) | 396404 (45917) | 244672 (161520) | 72119 (311554) | 1 (367391) |
| *PALB2* het without p.Trp1038Ter | 498 (0) | 496 (0) | 473 (0) | 397 (47) | 227 (156) | 68 (287) | 0 (336) |
| *PALB2* p.Trp1038Ter | 312 (0) | 314 (0) | 292 (0) | 241 (24) | 128 (102) | 39 (181) | 0 (207) |

B.

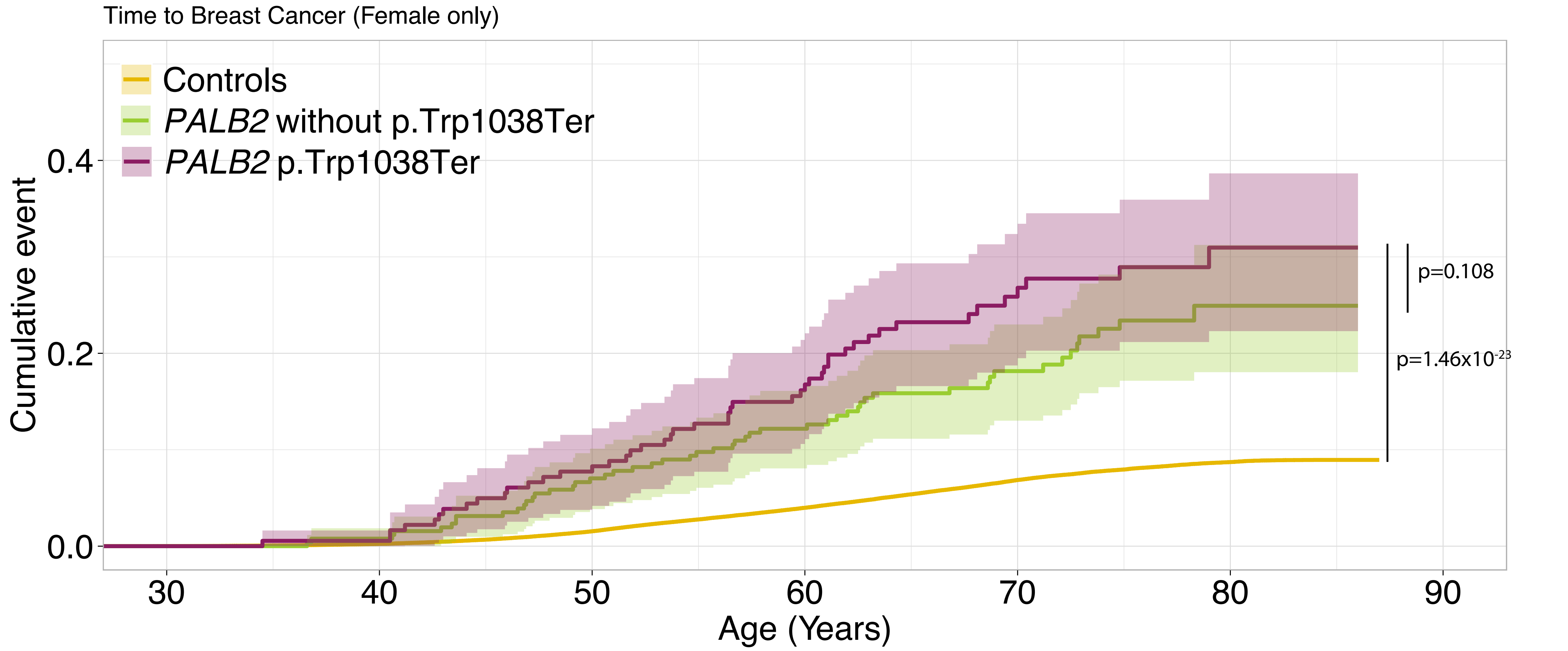


Number at risk (number censored)

| | | | | | | | |
|---|---|---|---|---|---|---|---|
| Controls | 237877 (0) | 253419 (0) | 250008 (151) | 224488 (25509) | 150427 (96124) | 50413 (196364) | 0 (236187) |
| *PALB2* het without p.Trp1038Ter | 256 (0) | 254 (0) | 238 (0) | 204 (27) | 135 (86) | 43 (173) | 0 (205) |
| *PALB2* p.Trp1038Ter | 181 (0) | 180 (0) | 167 (0) | 139 (15) | 80 (60) | 29 (116) | 0 (134) |
