## Supplemental Figure 11 for "Genomic ascertainment of *PALB2*-related cancer predisposition"

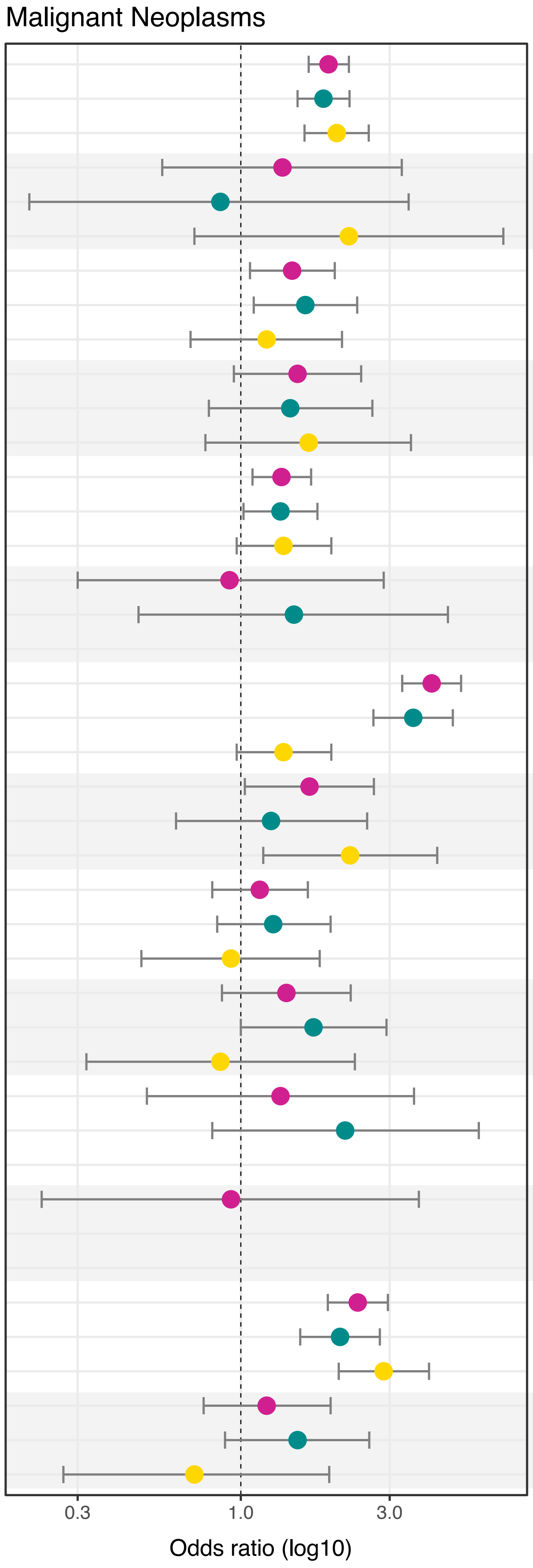


| | controls(%) | heterozygotes(%) | OR [95% CI] | p-value |
|---|---|---|---|---|
| All cancer | 115470(24.64) | 306(37.27) | 1.91[1.65–2.22] | $1.37\times10^{-17}$ |
| | 115470(24.64) | 185(36.85) | 1.84[1.52–2.23] | $3.00\times10^{-10}$ |
| | 115470(24.64) | 121(37.93) | 2.03[1.60–2.57] | $6.30\times10^{-9}$ |
| C00–C14:Malignant neoplasms of lip, oral cavity and pharynx | 2135(0.46) | 5(0.61) | 1.36[0.56–3.28] | 0.49 |
| | 2135(0.46) | 2(0.40) | 0.86[0.21–3.45] | 0.83 |
| | 2135(0.46) | 3(0.94) | 2.22[0.71–6.94] | 0.17 |
| C15–C26:Malignant neoplasms of digestive organs | 17076(3.64) | 42(5.12) | 1.46[1.07–2.00] | 0.02 |
| | 17076(3.64) | 29(5.78) | 1.61[1.10–2.36] | 0.01 |
| | 17076(3.64) | 13(4.08) | 1.21[0.69–2.11] | 0.51 |
| C30–C39:Malignant neoplasms of respiratory and intrathoracic organs | 7140(1.52) | 18(2.19) | 1.52[0.95–2.43] | 0.08 |
| | 7140(1.52) | 11(2.19) | 1.44[0.79–2.64] | 0.23 |
| | 7140(1.52) | 7(2.19) | 1.65[0.77–3.51] | 0.20 |
| C43–C44:Melanoma and other malignant neoplasms of skin | 43745(9.33) | 98(11.94) | 1.35[1.09–1.68] | 0.01 |
| | 43745(9.33) | 60(11.95) | 1.34[1.02–1.76] | 0.04 |
| | 43745(9.33) | 38(11.91) | 1.37[0.97–1.95] | 0.07 |
| C45–C49Malignant neoplasms of mesothelial and soft tissue | 1900(0.41) | 3(0.37) | 0.92[0.30–2.87] | 0.89 |
| | 1900(0.41) | 3(0.60) | 1.48[0.47–4.61] | 0.50 |
| | 1900(0.41) | 0(0) | NA | NA |
| C50:Malignant neoplasms of breast | 20171(4.3) | 112(13.64) | 4.09[3.29–5.08] | $6.21\times10^{-37}$ |
| | 20171(4.3) | 60(11.95) | 3.57[2.66–4.78] | $1.80\times10^{-17}$ |
| | 20171(4.3) | 38(11.91) | 1.37[0.97–1.95] | 0.07 |
| C51–C58:Malignant neoplasms of female genital organs | 6568(2.59) | 21(4.81) | 1.66[1.03–2.67] | 0.04 |
| | 6568(2.59) | 9(3.52) | 1.25[0.62–2.54] | 0.53 |
| | 6568(2.59) | 12(6.63) | 2.24[1.18–4.26] | 0.01 |
| C60–C63:Malignant neoplasms of male genital organs | 17839(8.31) | 35(9.11) | 1.15[0.81–1.64] | 0.43 |
| | 17839(8.31) | 25(10.16) | 1.27[0.84–1.94] | 0.26 |
| | 17839(8.31) | 10(7.25) | 0.93[0.48–1.79] | 0.83 |
| C64–C68:Malignant neoplasms of urinary tract | 7453(1.59) | 18(2.19) | 1.40[0.87–2.25] | 0.16 |
| | 7453(1.59) | 14(2.79) | 1.71[1.00–2.93] | 0.05 |
| | 7453(1.59) | 4(1.25) | 0.86[0.32–2.32] | 0.76 |
| C69–C72:Malignant neoplasms of eye, brain and other parts of central nervous system | 1726(0.37) | 4(0.49) | 1.34[0.50–3.59] | 0.56 |
| | 1726(0.37) | 4(0.80) | 2.16[0.81–5.79] | 0.13 |
| | 1726(0.37) | 0(0) | NA | NA |
| C73–C75:Malignant neoplasms of thyroid and other endocrine glands | 1245(0.27) | 2(0.24) | 0.93[0.23–3.72] | 0.91 |
| | 1245(0.27) | 1(0.20) | NA | NA |
| | 1245(0.27) | 1(0.31) | NA | NA |
| C76–C80:Malignant neoplasms of ill–defined, other secondary and unspecified sites | 24584(5.25) | 94(11.45) | 2.37[1.90–2.96] | $1.56\times10^{-14}$ |
| | 24584(5.25) | 52(10.36) | 2.08[1.55–2.79] | $1.02\times10^{-6}$ |
| | 24584(5.25) | 42(13.17) | 2.87[2.06–4.01] | $5.78\times10^{-10}$ |
| C81–C96:Malignant neoplasms of lymphoid, hematopoietic and related tissue | 8657(1.85) | 19(2.31) | 1.21[0.76–1.94] | 0.42 |
| | 8657(1.85) | 14(2/79) | 1.52[0.89–2.58] | 0.13 |
| | 8657(1.85) | 5(1.57) | 0.71[0.27–1.92] | 0.50 |
