## Supplemental Figure 7 for "Genomic ascertainment of *PALB2*-related cancer predisposition"

### *PALB2* Benign Neoplasms

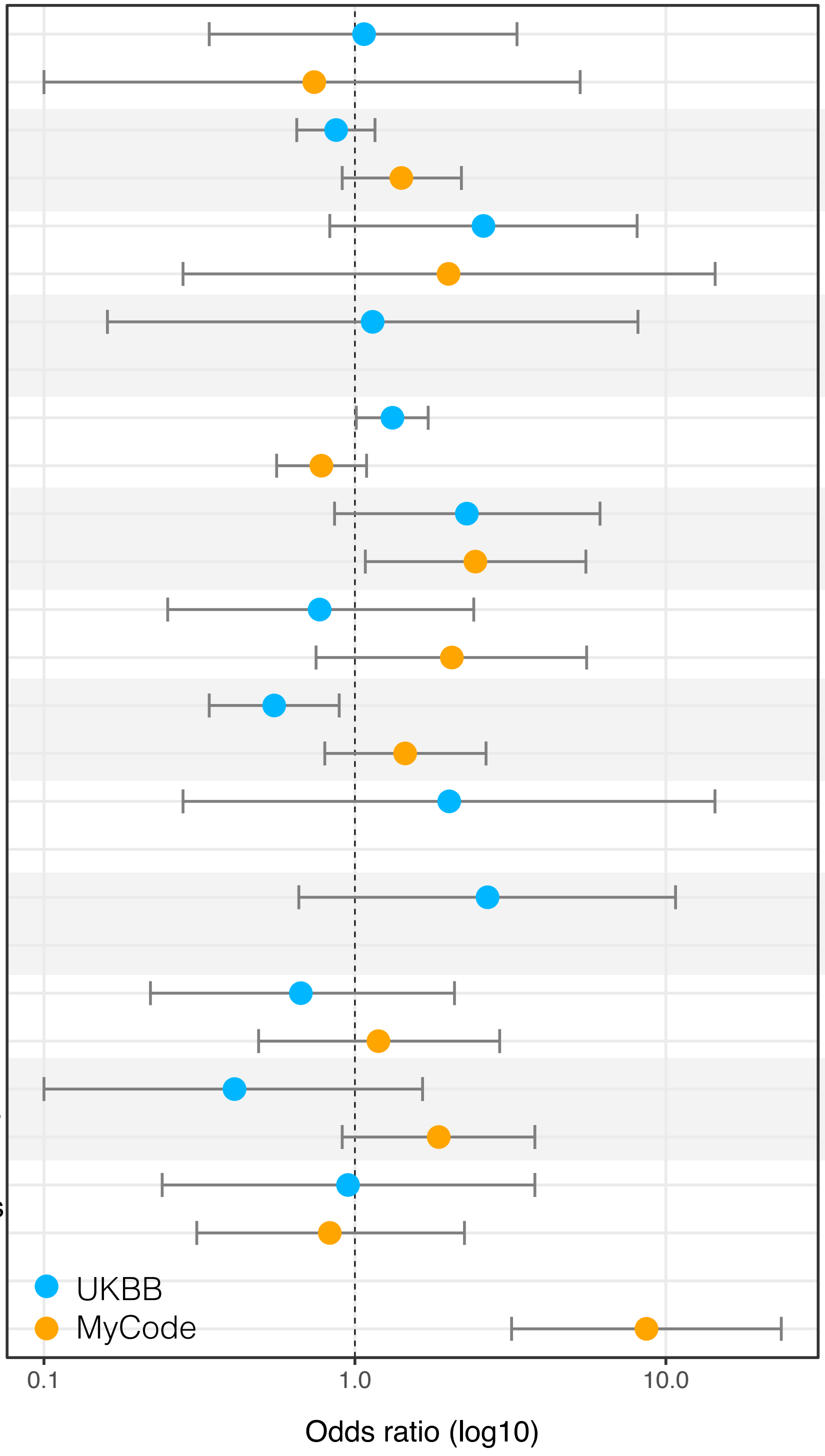


| | controls(%) | heterozygotes(%) | OR [95% CI] | p–value |
|---|---|---|---|---|
| D10–D11:Benign neoplasm of mouth, pharynx and major salivary glands | 1618(0.35) | 3(0.37) | 1.07[0.34–3.32] | 0.91 |
| | 1335(0.80) | 1(0.56) | 0.74[0.10–5.30] | 0.77 |
| D12–D13:Benign neoplasm of digestive organs | 33330(7.11) | 51(6.21) | 0.87[0.65–1.16] | 0.34 |
| | 19641(11.84) | 25(13.96) | 1.41[0.91–2.20] | 0.13 |
| D14–D15:Benign neoplasm of respiratory and intrathoracic organs | 664(0.14) | 3(0.37) | 2.59[0.83–8.08] | 0.10 |
| | 518(0.31) | 1(0.56) | 2.00[0.28–14.40] | 0.49 |
| D16:Benign neoplasm of bone and articular cartilage | 502(0.11) | 1(0.12) | 1.14[0.16–8.13] | 0.90 |
| | 804(0.48) | 0(0) | NA | NA |
| D17,D18,D22,D23:Benign neoplasms of skin | 25714(5.49) | 58(7.06) | 1.32[1.01–1.72] | 0.04 |
| | 53370(32.17) | 48(26.81) | 0.78[0.56–1.09] | 0.14 |
| D19–D21:Benign neoplasm of mesothelial and soft tissue | 1011(0.22) | 4(0.49) | 2.29[0.86–6.14] | 0.10 |
| | 2333(1.41) | 6(3.35) | 2.44[1.08–5.53] | 0.03 |
| D24:Benign neoplasm of breast | 2252(0.48) | 3(0.37) | 0.77[0.25–2.41] | 0.66 |
| | 1759(1.06) | 4(2.23) | 2.05[0.75–5.56] | 0.16 |
| D25–D27:Benign neoplasm of female genital organs | 17537(6.90) | 18(4.11) | 0.55[0.34–0.89] | 0.01 |
| | 7743(7.64) | 12(10.71) | 1.45[0.80–2.64] | 0.22 |
| D29:Benign neoplasm of male genital organs | 280(0.13) | 1(0.26) | 2.01[0.28–14.38] | 0.49 |
| | 391(0.61) | 0(0) | NA | NA |
| D30:Benign neoplasm of urinary organs | 431(0.09) | 2(0.24) | 2.67[0.66–10.74] | 0.17 |
| | 291(0.175) | 0(0) | NA | NA |
| D31–D33:Benign neoplasm of eye, brain, and other parts of CNS | 2593(0.55) | 3(0.37) | 0.67[0.22–2.09] | 0.49 |
| | 4212(2.539) | 5(2.79) | 1.19[0.49–2.92] | 0.70 |
| D34–D35:Benign neoplasm of endocrine organs | 2797(0.60) | 2(0.24) | 0.41[0.10–1.65] | 0.21 |
| | 4278(2.579) | 8(4.46) | 1.86[0.91–3.79] | 0.09 |
| D36:Benign neoplasm of other and unspecified sites | 1224(0.26) | 2(0.24) | 0.95[0.24–3.79] | 0.94 |
| | 4617(2.783) | 4(2.234) | 0.83[0.31–2.25] | 0.72 |
| D3A:Benign neuroendocrine tumors | 467(0.281) | 4(2.234) | 8.66[3.19–23.50] | $2.21\times10^{-5}$ |
