## Supplemental Figure 6 for "Genomic ascertainment of *PALB2*-related cancer predisposition"

*PALB2* Carcinoma in situ

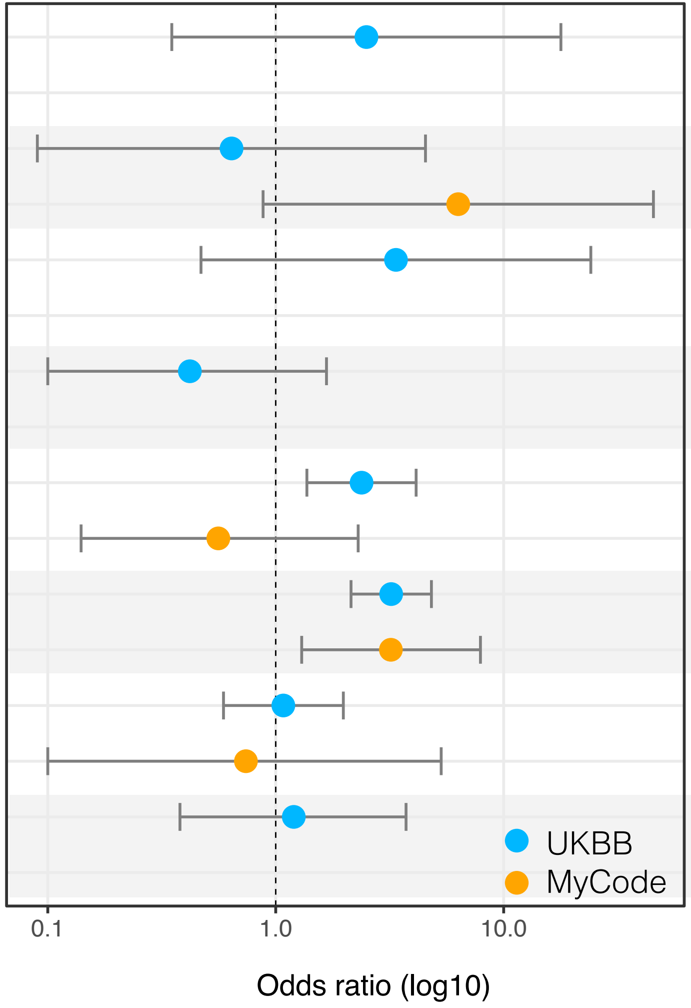


| | controls(%) | heterozygotes(%) | OR [95% CI] | p–value |
|---|---|---|---|---|
| D00:Carcinoma in situ of oral cavity, esophagus and stomach | 234(0.05) | 1(0.12) | 2.5[0.35–17.83] | 0.36 |
| | 52(0.031) | 0(0) | NA | NA |
| D01:Carcinoma in situ of other and unspecified digestive organs | 907(0.19) | 1(0.12) | 0.64[0.09–4.54] | 0.65 |
| | 163(0.098) | 1(0.558) | 6.32[0.88–45.4] | 0.07 |
| D02:Carcinoma in situ of middle ear and respiratory system | 176(0.04) | 1(0.12) | 3.37[0.47–24.12] | 0.23 |
| | 29(0.01) | 0(0) | NA | NA |
| D03:Melanoma in situ | 2788(0.59) | 2(0.24) | 0.42[0.1–1.67] | 0.22 |
| | 1341(0.808) | 0(0) | NA | NA |
| D04:Carcinoma in situ of skin | 3294(0.70) | 13(1.58) | 2.38[1.37–4.13] | $2.09 \times 10^{-3}$ |
| | 3770(2.273) | 2(1.117) | 0.56[0.14–2.3] | 0.42 |
| D05:Carcinoma in situ of breast | 4787(1.02) | 25(3.05) | 3.21[2.14–4.82] | $1.62 \times 10^{-8}$ |
| | 1633(0.984) | 5(2.793) | 3.2[1.3–7.91] | 0.01 |
| D06:Carcinoma in situ of cervix uteri | 5866(1.25) | 11(1.34) | 1.08[0.59–1.98] | 0.79 |
| | 1162(1.147) | 1(0.892) | 0.74[0.1–5.32] | 0.77 |
| D07:Carcinoma in situ of other_and unspecified genital organs | 1440(0.31) | 3(0.37) | 1.2[0.38–3.73] | 0.76 |
| | 466(0.28) | 0(0) | NA | NA |
