## Supplemental Figure 5 for "Genomic ascertainment of *PALB2*-related cancer predisposition"

### . *PALB2* Malignant Neoplasms

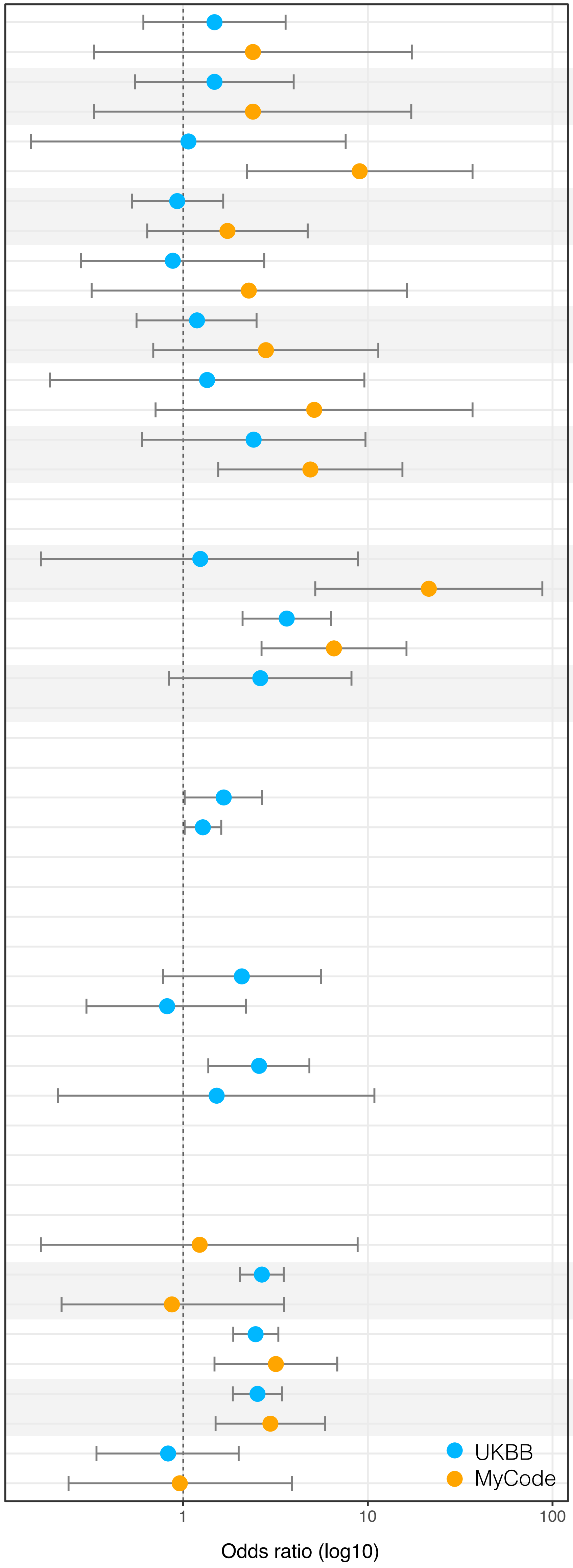


| | controls(%) | heterozygotes(%) | OR [95% CI] | p–value |
|---|---|---|---|---|
| C15:Malignant neoplasm of esophagus | 1944(0.41) | 5(0.61) | 1.48[0.61–3.59] | 0.38 |
| | 446(0.268) | 1(0.56) | 2.39[0.33–17.30] | 0.39 |
| C16:Malignant neoplasm of stomach | 1558(0.33) | 4(0.49) | 1.48[0.55–3.97] | 0.44 |
| | 441(0.27) | 1(0.56) | 2.39[0.33–17.20] | 0.39 |
| C17:Malignant neoplasm of small intestine | 543(0.12) | 1(0.12) | 1.07[0.15–7.59] | 0.95 |
| | 229(0.14) | 2(1.12) | 9.04[2.22–36.90] | $2.13 \times 10^{-3}$ |
| C18:Malignant neoplasm of colon | 7490(1.60) | 12(1.46) | 0.93[0.53–1.65] | 0.81 |
| | 2488(1.5) | 4(2.23) | 1.74[0.64–4.73] | 0.28 |
| C19:Malignant neoplasm of rectosigmoid junction | 1970(0.42) | 3(0.37) | 0.88[0.28–2.75] | 0.83 |
| | 455(0.274) | 1(0.56) | 2.27[0.32–16.30] | 0.41 |
| C20:Malignant neoplasm of rectum | 3429(0.73) | 7(0.85) | 1.19[0.56–2.50] | 0.65 |
| | 742(0.447) | 2(1.12) | 2.81[0.69–11.40] | 0.15 |
| C21:Malignant neoplasm of anus and anal canal | 436(0.09) | 1(0.12) | 1.35[0.19–9.59] | 0.77 |
| | 203(0.12) | 1(0.56) | 5.13[0.71–36.90] | 0.1 |
| C22:Malignant neoplasm of liver and intrahepatic bile ducts | 497(0.20) | 2(0.46) | 2.41[0.60–9.72] | 0.22 |
| | 652(0.393) | 3(1.68) | 4.89[1.55–15.40] | 0.01 |
| C23:Malignant neoplasm of gallbladder | 67(0.03) | 0(0) | NA. | NA. |
| | 46(0.03) | 0(0) | NA. | NA. |
| C24:Malignant neoplasm of other and unspecified parts_of biliary tract | 468(0.10) | 1(0.12) | 1.24[0.17–8.85] | 0.83 |
| | 101(0.06) | 2(1.12) | 21.40[5.20–88.10] | $2.22 \times 10^{-5}$ |
| C25:Malignant neoplasm of pancreas | 2113(0.45) | 13(1.58) | 3.64[2.10–6.32] | $4.38 \times 10^{-6}$ |
| | 827(0.498) | 5(2.79) | 6.56[2.66–16.2] | $4.33 \times 10^{-5}$ |
| C26:Malignant neoplasm of other and ill–defined digestive organs | 668(0.14) | 3(0.37) | 2.62[0.84–8.16] | 0.1 |
| | 60(0.036) | 0(0) | NA. | NA. |
| C43:Melanoma | 6004(1.28) | 17(2.07) | 1.66[1.02–2.68] | 0.04 |
| C44:Other malignant neoplasms of skin | 39738(8.48) | 85(10.35) | 1.28[1.02–1.61] | 0.04 |
| C51:Malignant neoplasm of vulva | 292(0.11) | 0(0) | NA. | NA. |
| C52:Malignant neoplasm of vagina | 78(0.03) | 1(0.22) | NA. | 0.97 |
| C53:Malignant neoplasm of cervix uteri | 1143(0.45) | 4(0.92) | 2.08[0.78–5.59] | 0.14 |
| C54:Malignant neoplasm of corpus uteri | 2894(1.14) | 5(1.14) | 0.82[0.3–2.19] | 0.69 |
| C55:Malignant neoplasm of uterus, part unspecified | 335(0.13) | 0(0) | NA. | NA. |
| C56:Malignant neoplasm of ovary | 2333(0.92) | 12(2.75) | 2.58[1.37–4.83] | $3.19 \times 10^{-3}$ |
| C57:Malignant neoplasm of other and unspecified female genetial organs | 392(0.15) | 1(0.23) | 1.52[0.21–10.86] | 0.68 |
| C58:Malignant neoplasm of placenta | 25(0.01) | 0(0) | NA. | NA. |
| C76:Malignant neoplasm of other and ill–defined sites | 319(0.07) | 0(0) | NA. | NA. |
| | 829(0.499) | 1(0.558) | 1.23[0.17–8.81] | 0.84 |
| C77:Secondary and unspecified malignant neoplasm of lymph nodes | 12837(2.74) | 57(6.94) | 2.67[2.03–3.51] | $1.84 \times 10^{-12}$ |
| | 2305(1.389) | 2(1.117) | 0.87[0.22–3.53] | 0.85 |
| C78:Secondary malignant neoplasm of respiratory and digestive_organs | 13127(2.80) | 54(6.58) | 2.47[1.87–3.28] | $2.86 \times 10^{-10}$ |
| | 2362(1.424) | 7(3.91) | 3.18[1.48–6.84] | $3.00 \times 10^{-3}$ |
| C79:Secondary malignant neoplasm of other and unspecified sites | 10596(2.26) | 44(5.36) | 2.53[1.86–3.43] | $3.35 \times 10^{-9}$ |
| | 3340(2.013) | 9(5.027) | 2.97[1.5–5.88] | $1.81 \times 10^{-3}$ |
| C80:Malignant neoplasm without specification of site | 3525(0.75) | 5(0.61) | 0.83[0.34–2.00] | 0.67 |
| | 2148(1.29) | 2(1.117) | 0.96[0.24–3.89] | 0.96 |
